# Appraising Cardiovascular 10-yr Risk Prediction Scores: A Rapid Systematic Review

**DOI:** 10.1101/2025.02.07.25321756

**Authors:** Chiranjivi Adhikari, Komal Shah, Aakansha Shukla, Biraj Man Karmacharya, Dileep Mavalankar

## Abstract

Globally, the burden of cardiovascular disease is on the rise. Despite WHO’s and the UN’s frantic efforts, it appears less probable that the 25 by 25 aim will be met. Early identification of at-risk cases using a risk scoring system can aid in achieving these goals, however for primary and secondary prevention, suitability of these scoring systems, for the countries with medium to low resources, including Asians, with respect to accuracies is a challenge as majority of them are developed from non-Asian cohorts. In light of methodological considerations, risk attribution, and policy consequences, we included and described, restricting our search but sytematically, with five widely used global tools for CVD risk 10-year prediction—FRS, WHO CVD, QRISK, ASCVD, and SCORE—and their updated versions, altogether 11, published during 1970-2023. In general, the results of consolidated risk ratings and summarization showed that these algorithms can differentiate CVD 10-yr risk by 63-86% accurately, considering both for internal and external validity. Further, we discuss their methodological perspectives, ad hoc use, and suggest prospects.

**Registration:** Open Science Framework (https://osf.io/72v48)

**Highlights:**

- Globally and optimally used cardiovascular disease (CVD) risk scoring algorithms have diverse accuracies, generalizability, and levels of evidence based on study designs, analyses, and given gold standards.
- Although validated fair to excellent, still the performance of these tools can be increased from 12 to 37%, both for internal and external accuracy, which can have a positive impact on cost and public health.
- Lifestyle and related changes due to gene-environment interaction, these algorithms are liable to change, so we need to update, validate, and fit them accordingly.
- Implementational, methodological, technological, and cost-related issues need to be addressed for a country or a state-specific algorithm to be up taken, updated, or validated.

## Background

Global burden of disease 2017 Causes of Death Collaborators and a report from the American Heart Association (AHA) reveal that cardiovascular diseases (CVD) share nearly one-third (> 17.3 million, 32%) of all-cause mortality, and from almost two-thirds to a half (42-50 %) of total noncommunicable disease deaths, which increased by 21 and 42 percentage points in 2007-2017 and 1990-2013 [1–3], respectively. The death toll is expected to reach 23.6 million by 2030 [1] and remains the leading killer and morbidity cause, globally [1, 2]. As the World Health Organization (WHO) proposes an ambition of 25% reduction in noncommunicable diseases (NCDs) by 2025 [3], there must be at least two-fifths decrease in CVD mortality as par its proportion in NCD deaths. Furthermore, more than half of the world’s countries are failing to meet the 2030 agenda for sustainable development goal (SDG) target of reducing premature deaths from NCDs by one-third (of 2015 status) [4]. Moreover, the estimated global cost of CVD is expected to increase from 863 in 2010 to 1044 billion (US $) in 2030 [5] and this mammoth of a health care burden imparted on the global economy has necessitated several efforts towards the design and implementation of strategies to improve the diagnosis, treatment, and prognosis of CVDs. Furthermore, 70 to 80% of those deaths occur in low-and-middle-income-countries (LMICs) [6, 7]. With the chronic nature of the disease and recent advances in health care innovations, it is possible to improve life expectancy of the population living with CVD and this large proportion of CVD can be prevented by aggressive risk factor modification in the individuals who are at risk of developing the disease.

Cardiovascular diseases are multifactorial, and disparities in these factors underlie in the socio-demographic and geographic backgrounds of the populations [8]. Accordingly, identifying individuals at the highest risk of developing CVD and targeting appropriate prevention and treatment strategies have been the focus of research for over a couple of decades. For this, a surveillance system, mainly covering the preventive aspects, is needed, provided that policy favored and economically sound risk scoring tools are in place [7,9]. To predict the disease risk and quality of life before (primary prevention) and after the primary event of the CVD (secondary and tertiary prevention), several risk scoring algorithms have been developed in the last several decades, which principally estimate the short-and long-term risks of CVD events. These algorithms have been developed to calculate a CVD event risk for an individual over a 5/ 10-year period, and a few of them to calculate the lifetime risk, after identifying the statistically weighted contribution of modifiable and non-modifiable risk factors, the laboratory and nonlaboratory parameters.

The CVD risk algorithms are modelled, calibrated, and validated through representative samples, using multivariable strategies with large observational (mainly cohort) studies representing ethnicities, or geographic boundaries, and are liable to inherent error and variability due to settings, population, emergence of newer risk factors, and local or national epidemiological shifts in the disease characteristics. Even though the performances of these tools are found satisfactory to excellent, those factors implicate the discrepancies within and between internal and external validity and calibration scores, and so, creating a difficulty to discern in health and related policies, and implementation in decision-making, mainly for LMICs and Asian populations. Similarly, some of these algorithms may be either costly because more advanced tests are inherently needed or their cost-related analyses, including health technology assessments (HTAs), have never been performed, or they are only applicable at hospital and referral level facilities rather than primary care levels, where we can carry out screening and other cost-minimization techniques for resource-low settings. So, we aimed to discern the answers from global perspectives of the following research question: What are the consolidated range values of internal and external validity and methodological perspectives of CVD 10-yr risk scores?

### Evidence gaps

Globally used algorithms are neither originally developed nor the calibrated ones among Asian populations show consistencies with the original ones [10–13]. Even worse is the emerging public health problem that native and migrant South Asians are at increased CVD risk compared to the rest of the world’s population [14], and either appropriately calibrated current risk algorithms or ideally developed new ones are urgently needed [15].

## Methods

### Selection of risk scores, search strategy

We registered the protocol in the Open Science Framework (https://osf.io/72v48), bearing the research question in mind, restricting to five globally optimally used scoring algorithms— Framingham Risk Score (FRS), WHO CVD Risk 2019 (WHO 2019), Atherosclerotic Cardiovascular Disease (ASCVD), Systematic Coronary Risk Evaluation (SCORE), and QRESEARCH Cohort Database (QRISK) (ESM1.Tab1). CA carried out a search (validation AND cardiovascular AND (Framingham OR coronary risk OR world health OR qrisk OR atherosclerotic) with selective and parsimonious iterations, in the Google Scholar, PubMed and Hinari databases, followed by citation search in targeted cardiovascular related journals. Another review author (KS) scrutinized and endorsed to include eleven risk scores published during 1970-2023, including those derived from the original studies, for the review (Fig 1.).

**Fig 1.**
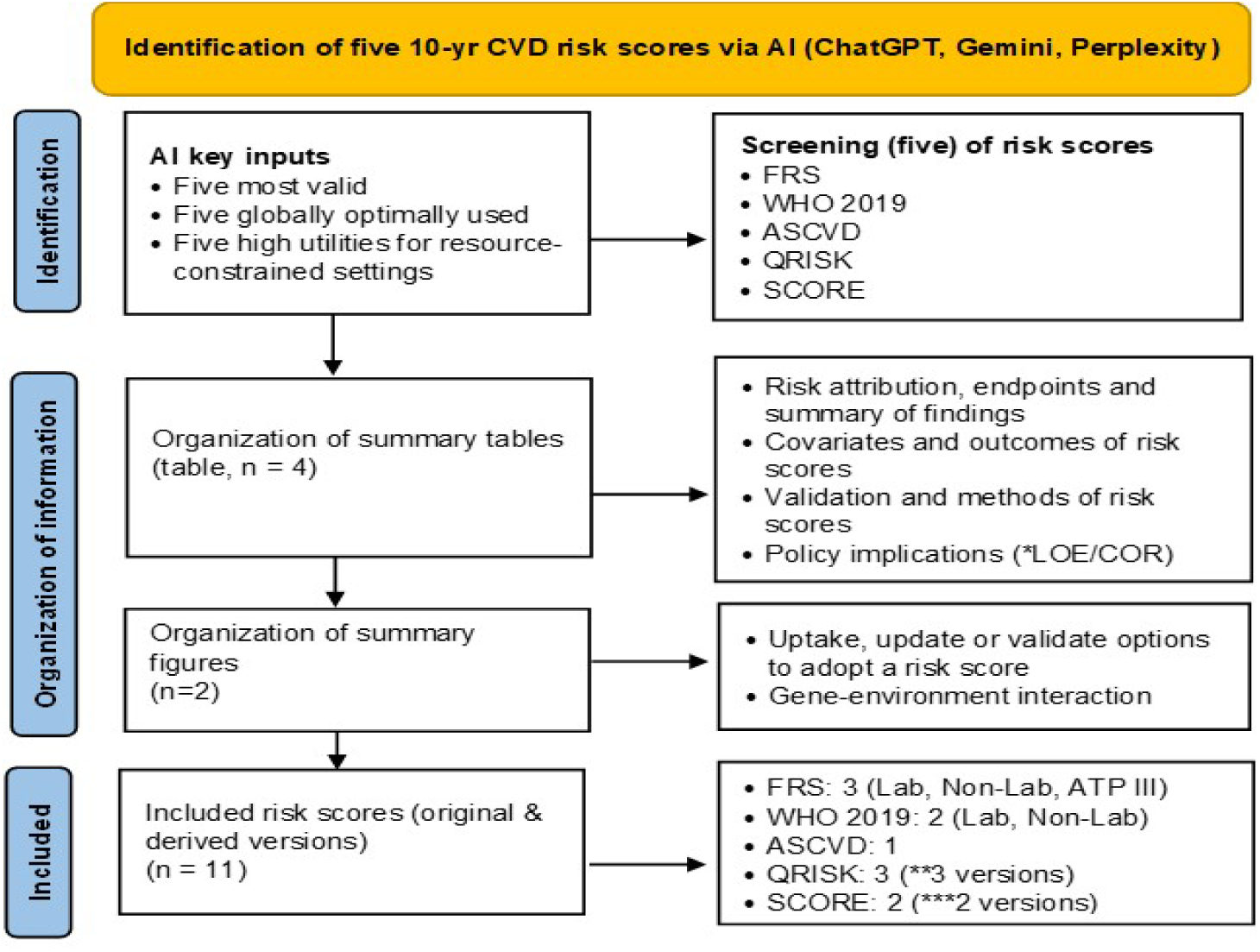
Inclusion of risk scores and studies, and information organization. *LOE/COR level of evidence/class of recommendation: **Includes QRISK, QRISK2 and QRISK3: ***Includes SCORE2 and SCORE2-OP

### Data extraction, summary and synthesis

We narrated methodological, parameter and outcome information, and policy-implicative information in three tables, and quantitatively consolidated the range values of C-statistics and additional information in the fourth one. Additionally, we conceptualized and proposed an appropriate selection and application pathway of a risk score tool to be needed and adopted by a state or a nation in a simple and understandable way, along with simpler explanation of the possible gene-environment interaction, in two different flow diagrams, respectively (Fig 1.). Since this is a review, ethical approval was not needed.

### Assessment of evidence and recommendation of risk scores

We summarized and followed the methods for the policy implicating attributes of the risk scores as recommended in literature and from authors’ experiences. Two authors (CA and AS), following AHA/ACC 2019 guideline [16], assessed the level (quality) of evidence (LOE) with categories A (high), B (moderate), and C (limited data and expert opinion), and the class (strength) of recommendation (COR) with categories I (strong), IIa (moderate), and IIb (weak). In case of discrepancies, both authors reached consensus.

### Assessment of performance metrics of risk scores

We found two performance indicators, validation and calibration, to evaluate the internal and external performances of a risk score, making four different indicators–internal validation and calibration for internal performance, and external validation (or transporting) and calibration (or recalibration in some literature) for external performance. For validation, we found most frequently used are: measures of discrimination, which include the area under the receiver operating characteristic curve (AUC-ROC) or C-statistics, or its pooled values (pooled C-statistics from meta-analysis), and concordance index (Harrel’s c-statistics or c-index), both similar in values and interpretation, however, the former is a function of sensitivity and specificity whereas the latter is a modification of C-statistics, which also accounts the model’s ability to predict risk at different levels. C-or c-statistics range between 0.5-1.0, and are interpreted as poor (0.5-0.6), fair (0.6-0.7), good (0.7-0.8), excellent (0.8-0.9), and outstanding (0.9-1.0). Less frequently used statistics are–correlation (r or ρ), sensitivity, specificity, accuracy, the Youden’s index, variance of the regressors (R^2^), Royston and Sauerbrei’s D-index (hereafter D-index), and variance of D-index (R-squared D, R^2^ ). The D-index measures the amount of variation in risk between individuals with low and high predicted risks, and may range between -∞ and +∞, whereas 0.5 or less indicates by chance, 0.6 as good, 0.7 and above as excellent, a negative score indicates worse, and the difference of scores of 0.1 is an indicator of an improved discriminatory outcome [17]. R^2^ is a modified version of traditional R^2^ statistics that adjusts the number of predictors in the model. The value ranges between 0 and 1, and generally interpreted as poor, moderate, good, and excellent for the values 0-0.1, 0.1-0.3, 0.3-0.5, and >0.5, respectively. For calibration, we compared quintiles or deciles of predicted vs observed datasets, Bald-Altman plot, calibration slope, Hosmer-Lemeshow χ^2^ goodness-of-fit statistics (H-L test), agreement percentage, and kappa value. Since there was variation in types of calibration measures used, we summarized benchmarking against kappa values and interpreted them as <0 (poor or by chance), 0.0-0.2 (slight), 0.2-0.4 (fair), 0.4-0.6 (moderate), 0.6-0.8 (substantial), and 0.8-1.0 (almost perfect) [18] or good-fit and poor-fit, in the H-L test and similar qualitative measurements, and in the authors’ consensuses.

For prospects, we explore the recalibration methods, which reevaluate the algorithms and include newer risk factors in a model, selected from hyperparameter tuning, where the indices like net reclassification index (NRI) and integrated discrimination improvement (IDI) are evaluated (Fig 2), which are described elsewhere [19].

**Fig 2.**
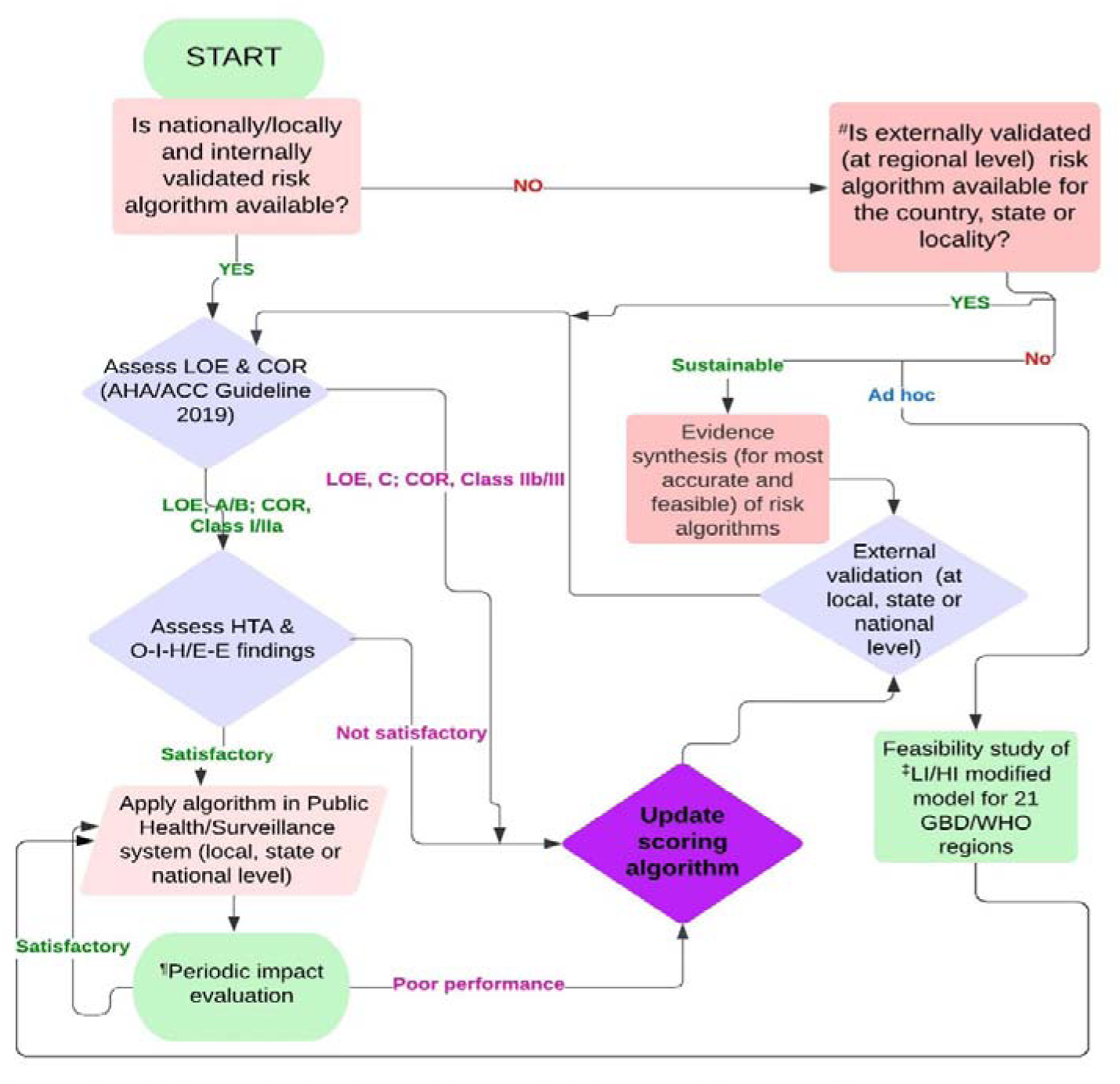
Flow diagram to uptake, update or validate a CVD risk score. ^#^Cautiously applied aligning with experts’ judgement when a regionally validated algorithm used at local, state or national level; ^‡^WHO/GBD 21 region-specifiec charts further derived model by Raghu et al. (https://doi.org/10.1371/journal.pone.0133618); ^¶^Period depends upon prediction duration, demographic, epidemiological or policy-related factors; Acronyms: AHA/ACC, American Heart Association/American College of Cardiology; LOE, Level (Quality) of Evidence[Categories, A, High, B, Moderate, C (includes C-LD, Limited Data, C-EO, Expert Opinion]; COR, Class (strength) of Recommendation [Categories. Class I, Strong, Class Ila. Moderate. Class lib, Weak, Class III, No Benefit, and Harm]; LOE and COR are independent to each other; HTA, Health Technology Assessment (or Equity/Economic Evaluation); O-l-H, Operational-Implementation-Health systems research continuum; E-E, Efficacy-Effectiveness research continuum; LI, Low Information (without Lipid); HI, High Information (with Lipid)

## Results

Globally used risk scores and their summary features, involved covariates, endpoints, their risk categories are depicted in table 1.

**Table 1.**
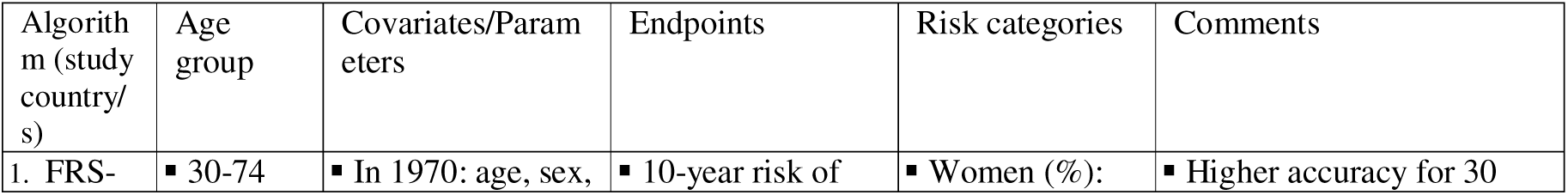
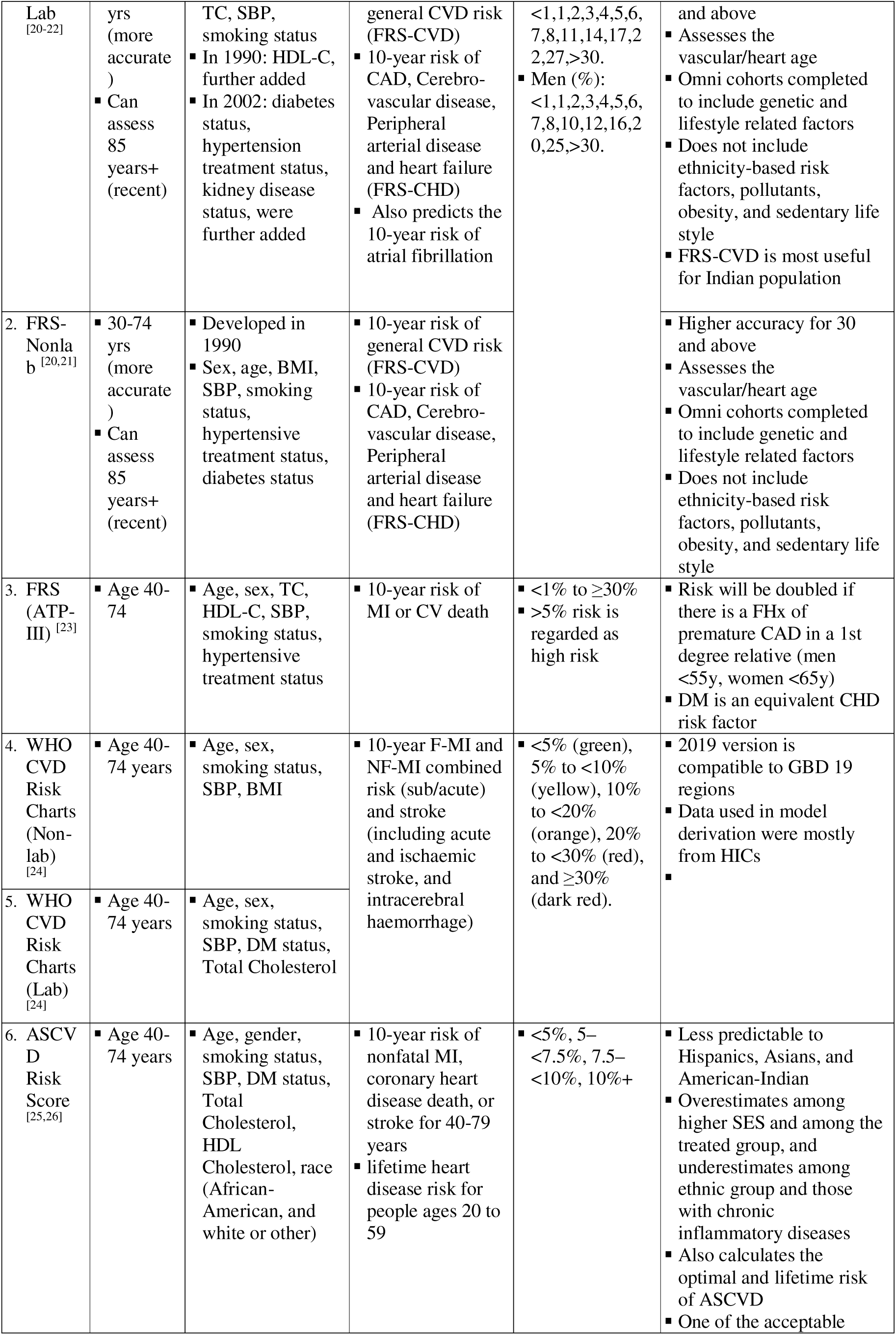
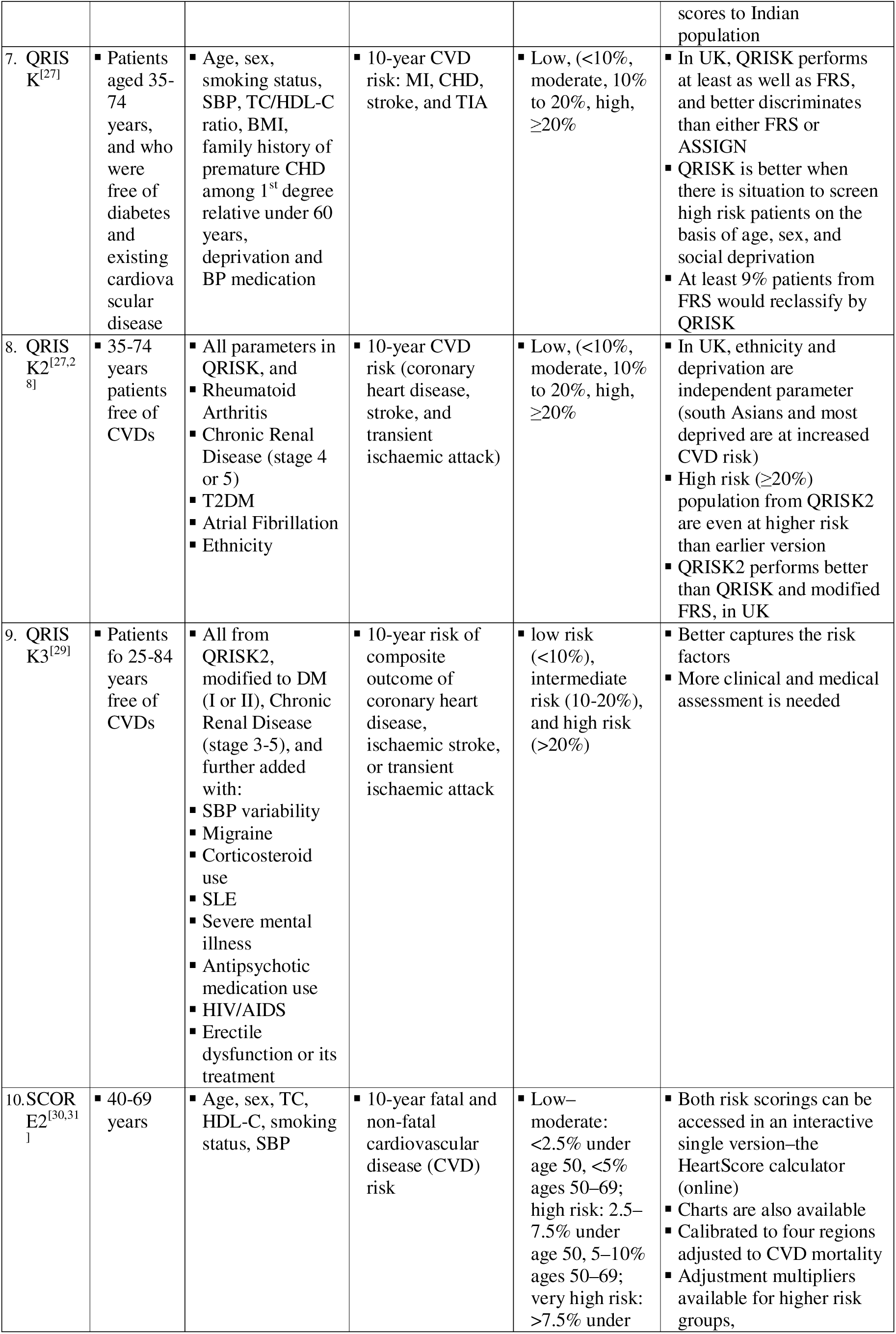
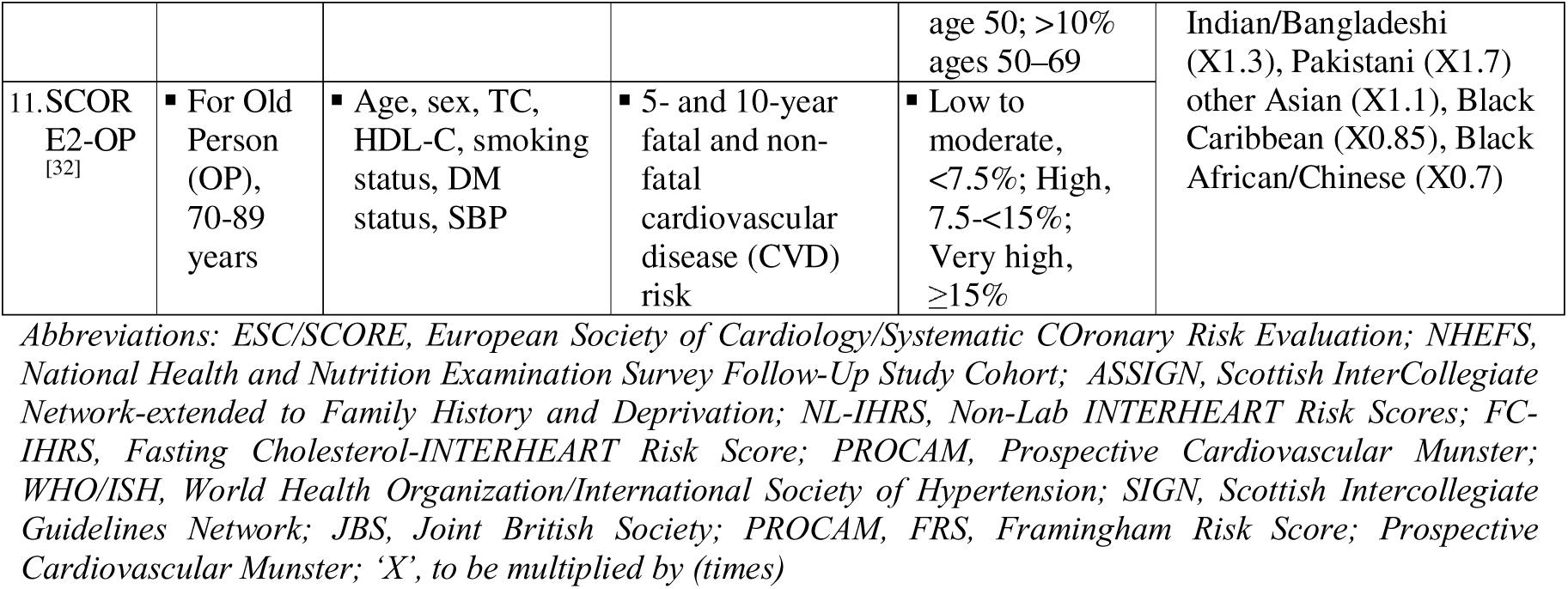
Risk attribution summary of algorithms and their updated versions.

Given the snapshot of covariates and outcomes, FRS Non-lab and WHO Non-lab are not based on laboratory testing. Age, sex, diastolic blood pressure, and body mass index are frequently used as covariates in non-laboratory risk scores. TC, HDL-C, and DM status, or blood sugar levels, are the common covariates in laboratory-based risk scores. Moreover, age, sex, and smoking status are the factors commonly included in both. Regarding outcomes, stroke, CHD death, and nonfatal MI are the frequent outcomes of risk scores, whereas fatal MI and CAD are the least frequent ones (Table 2).

**Table 2.**
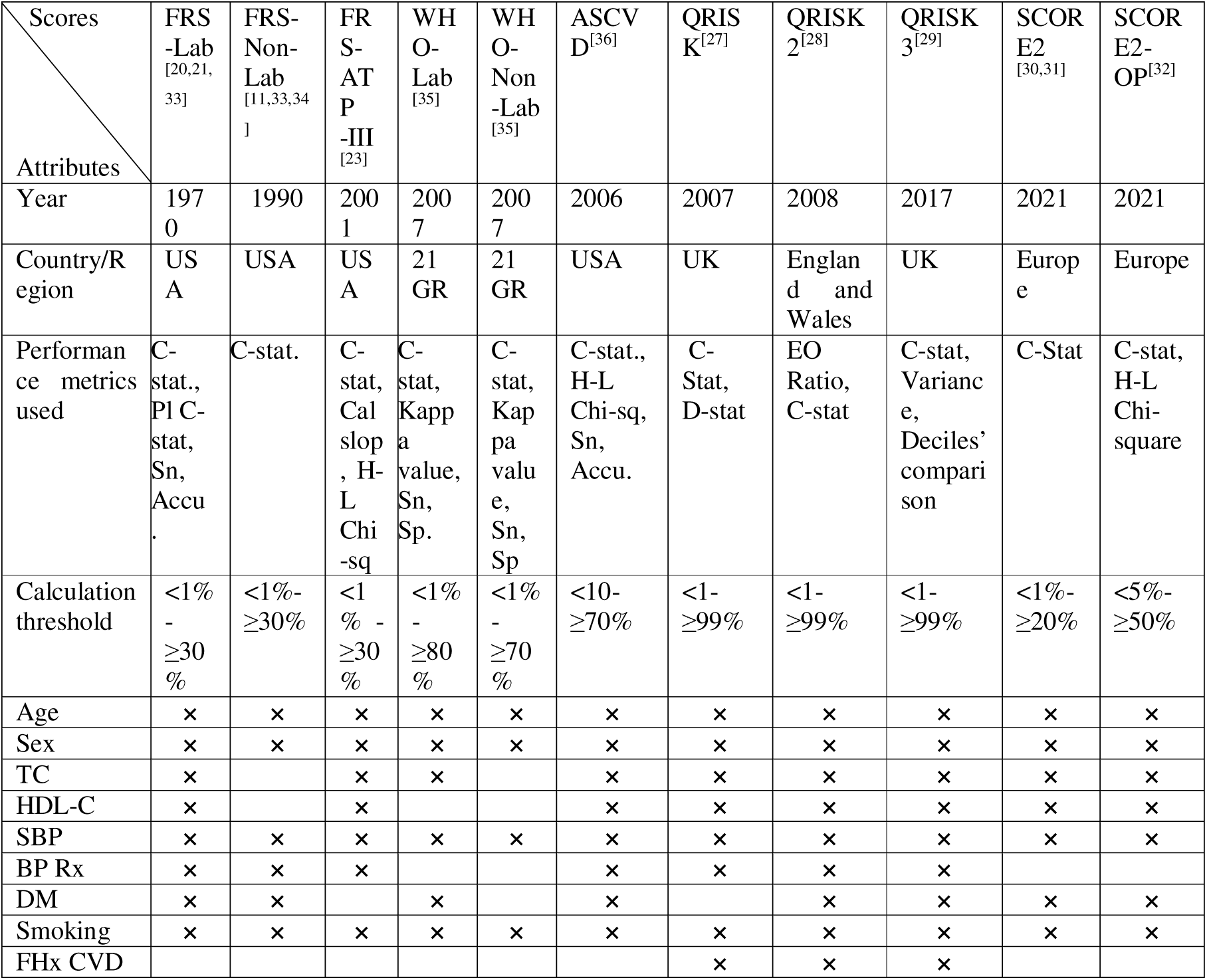
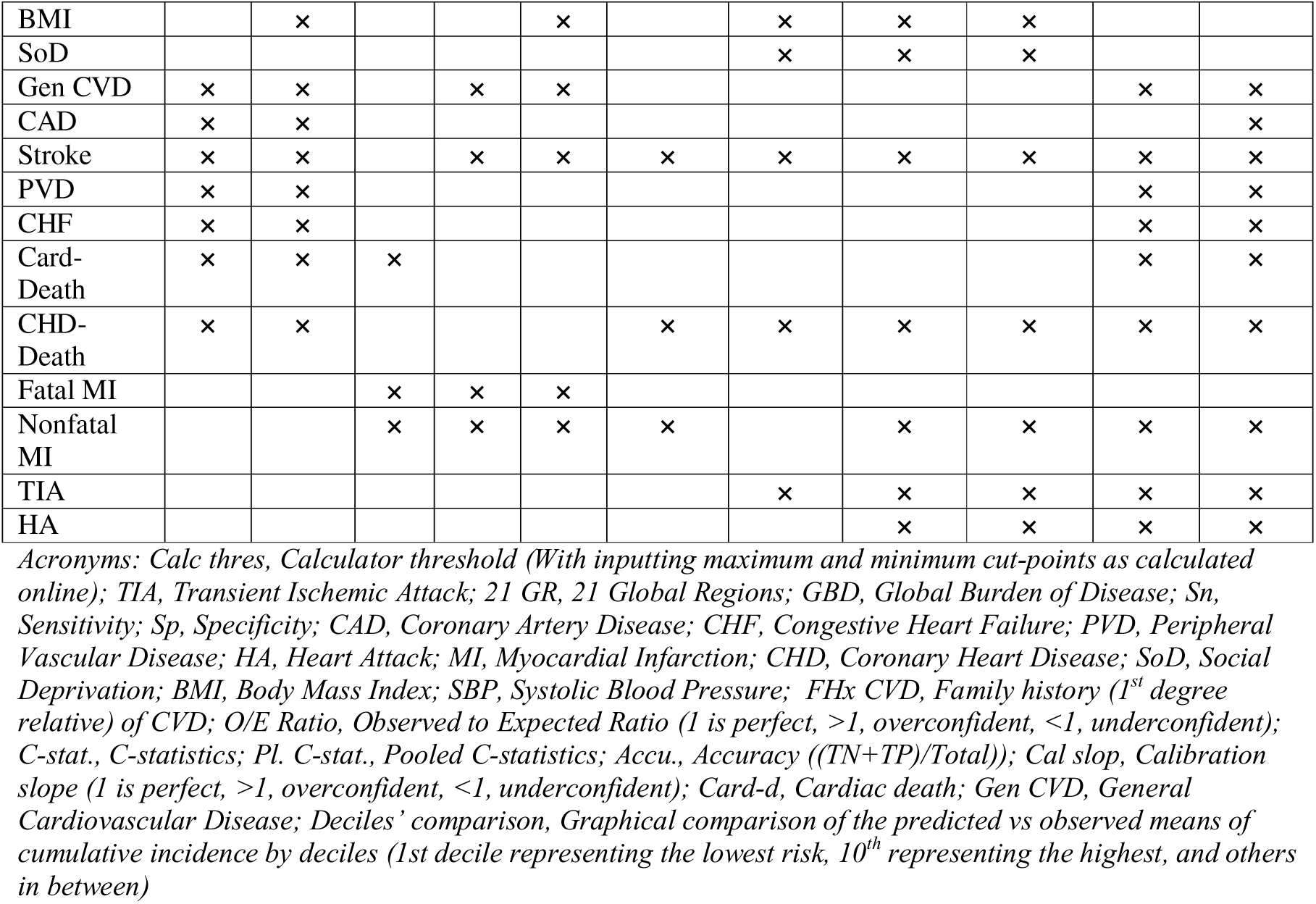
Covariates and outcomes of globally optimally used risk scores.

## Globally used scores for CVD 10-yr risk prediction

### Framingham Risk Score (FRS)

A classical risk scoring method was developed from the Framingham cohort established during the Framingham Heart Study of Boston, which started in 1948. The covariates inputted into the model in 1970 were age, sex, TC, SBP, and smoking status, whereas updates with HDL-C in 1990, and DM, BP treatment status, and the chronic kidney disease (CKD) were added in 2002 (Tables 1, 2). The internal and external validities remained at 0.72 and higher for both laboratory and non-laboratory scores (Table 3) [11,20,21,34,37]. The prime aim of this sex-specific multivariable risk factor algorithm was to identify the population at risk for developing CHD that can be hugely benefited by pharmacological interventions targeting high-risk and pharmacological as well as non-pharmacological interventions for individuals at intermediate risk, mainly to control hypertension and dyslipidemia. A cost-effective and sensitivity analysis showed that FRS non-laboratory parameters are equally sensitive, as validated with FRS and other lab-based risk scores [20,51] and 25-75% cost-effective when stage-based screening is carried out [51].

**Table 3.**
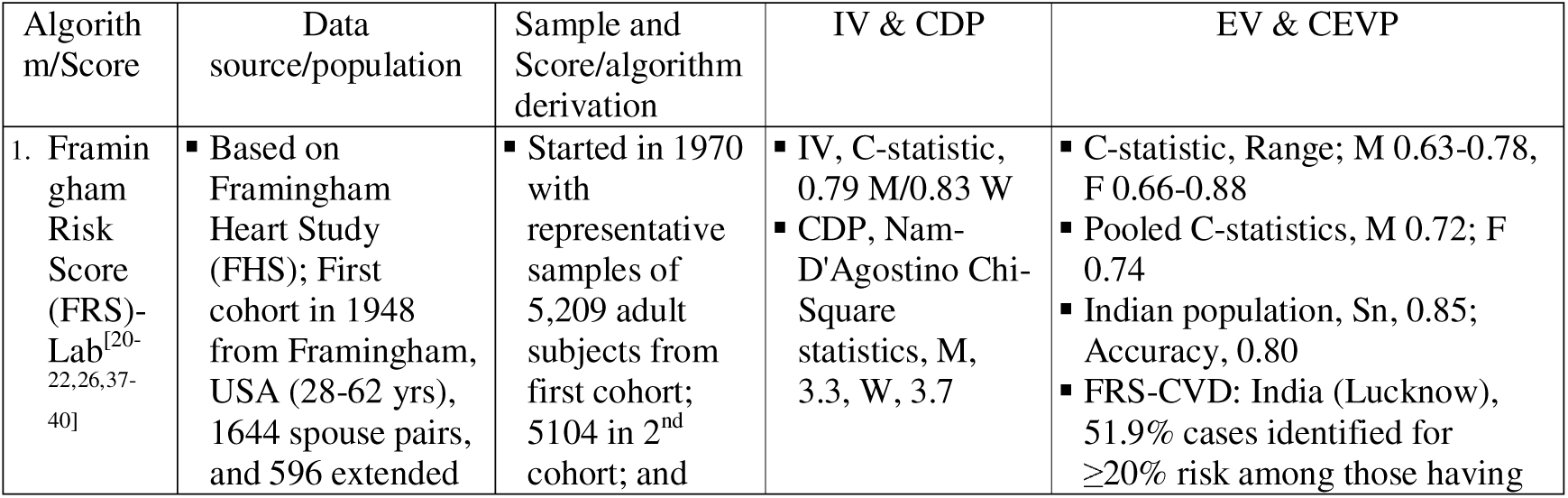
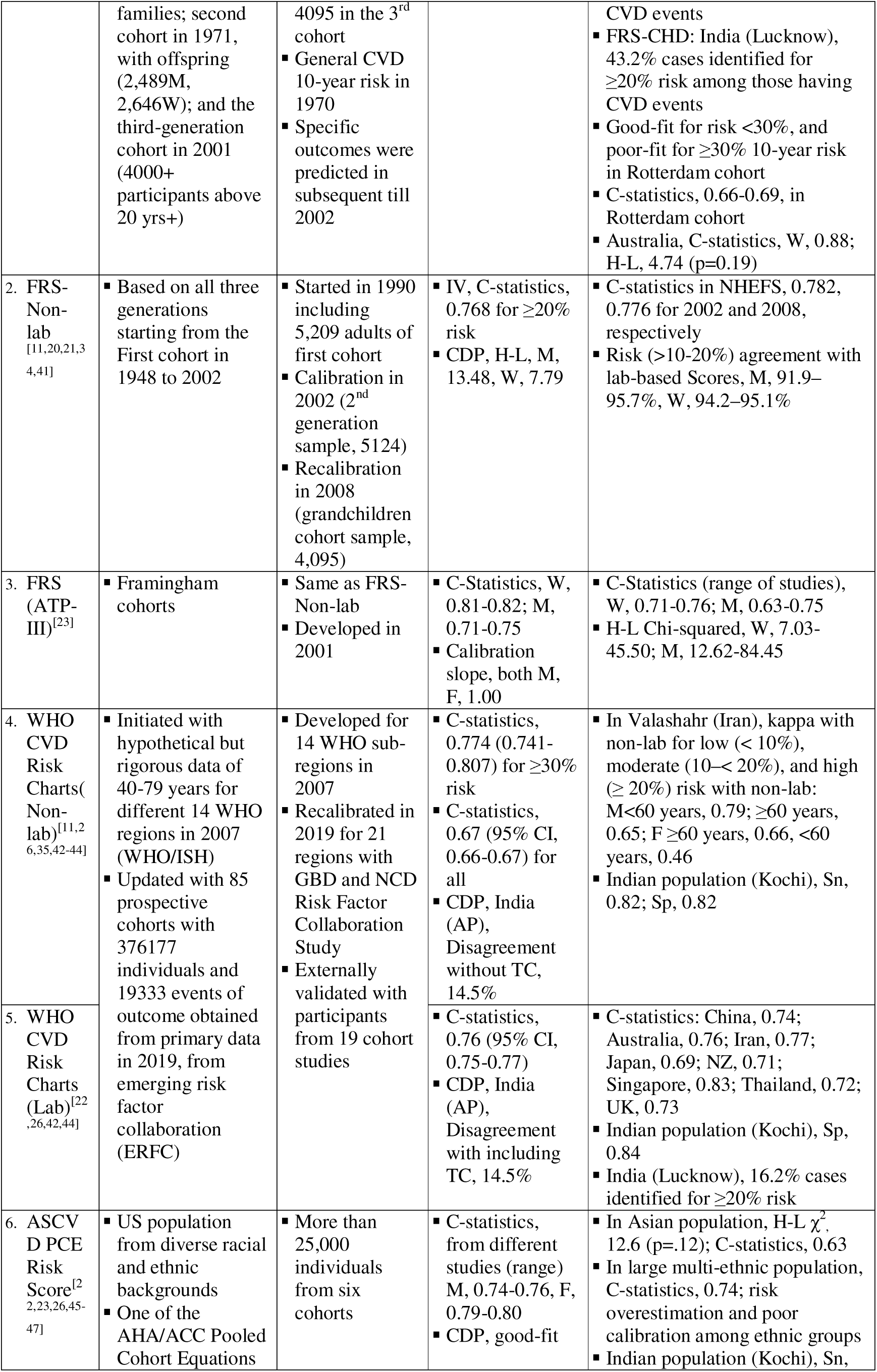
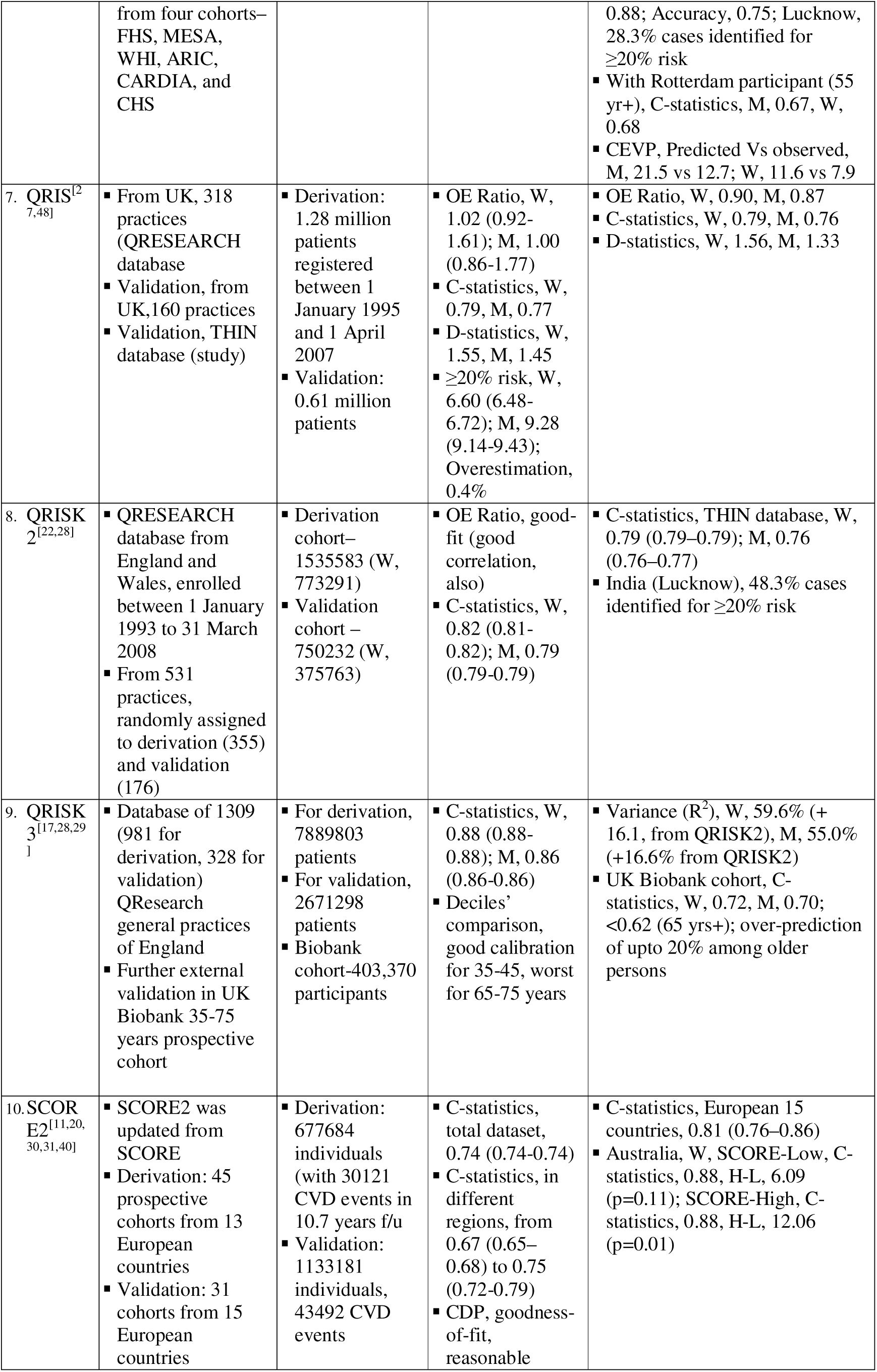
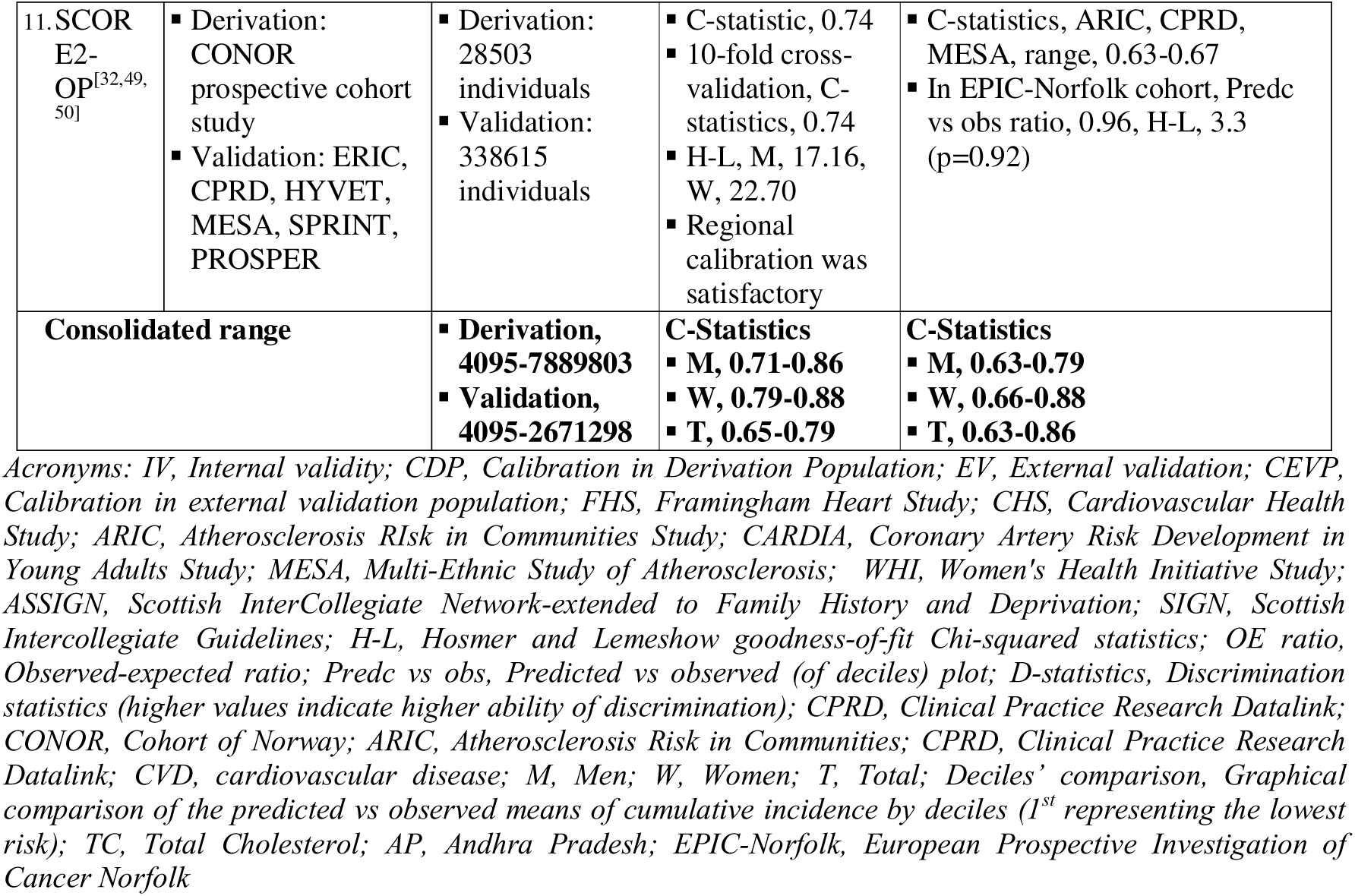
Method and validation summary of risk scores and their updated versions.

The first version of the risk algorithm was published in 1970 and laid the foundation stone for CV risk prediction [52]. The researchers found that Joint National Committee (JNC-V) blood pressure and National Cholesterol Education Program (NCEP) cholesterol guidelines of blood pressure, total cholesterol, and LDL cholesterol effectively predict coronary heart disease (CHD) risk in a middle-aged white population sample. A simple coronary disease prediction equation was developed using categorical variables, with the aim equipping healthcare providers to predict multivariate CHD risk in patients without overt CHD. It estimates 10-year risk for general and specific CVDs using various modifiable and non-modifiable determinants, namely age, sex, LDL cholesterol (LDL-C), HDL cholesterol (HDL-C), systolic blood pressure, and statuses of hypertension treatment, diabetes, and smoking, as laboratory parameters, and body mass index (BMI) in place of LDL-C and HDL-C, as nonlaboratory parameters. It classifies an individual in three categories: low risk (10% or less), intermediate risk (10-20%), and high risk (20% or more); however, the recently recalibrated algorithm yields the risks on an ordinal scale of <1% to >30%, giving further opportunities to analyze. Several recalibrations and external validations have been made to assess the appropriateness of the scoring in various ethnic groups, and it was found to perform fairly in American, European, and African races; however, for Asians, it was found to overestimate the risk [53] (Table 3).

As the original FRS only predicts 10-year risk of CHD, a modified algorithm was published in 2008 by ATP III (Adult Treatment Panel III), an expert panel of the National Heart, Lung, and Blood Institute (NIH, USA), to estimate a 10-yr risk of cardiovascular disease, cerebrovascular events, peripheral artery disease, and heart failure. The updated version was modified to include dyslipidemia, age range, hypertension treatment, smoking, and total cholesterol; and it excluded diabetes, because type 2 diabetes was considered to be a CHD risk equivalent, having the same 10-year risk as individuals with prior CHD [54]. In case of type 1 diabetes, a separate risk assessment system was considered with relatively less aggressive intervention goals. Moreover, in contrast to FRS-CHD, where coronary death and myocardial infarction were the only clinical endpoints, the updated FRS-CVD score included coronary insufficiency, angina pectoris, heart failure, TIA, peripheral arterial obstructive disease, stroke (ischemic and hemorrhagic), and cerebrovascular death as additional clinical endpoints. In the same domain as FRS risk scores, FRS (Adult Treatment Panel, ATP-III) was developed in 2001 to assess a 10-year risk of myocardial infarction (MI) or CV death, which has another important clinical significance in determining whether to initiate statins.

All FRS risk scores show their performances with both internally and externally valid, good to excellent, and calibrated good-fit. External validations were carried out in many countries and locations (Table 3). Though widespread, even the modified score suffers from a few limitations [55]. Cardiovascular diseases are multifactorial in nature with complex interactions with lifestyle, environmental, and genetic factors. In its two Omni cohorts, it has explored the genetic factors; however, both scoring algorithms have not addressed the ethnicity-based risk factors, pollutants, obesity (independent of BMI), and sedentary lifestyle (Table 1).

### WHO CVD Risk Charts

WHO developed, validated, both laboratory and non-laboratory cardiovascular risk charts, combining incidence from multiple sources and simulating a hypothetical 10-year risk from risk-factor distribution, relative risks, and CVD incidence in 2007 [24]. Keeping the same covariates for laboratory (age, sex, smoking status, SBP, DM status, and TC) and non-laboratory (age, sex, smoking status, SBP, and BMI), the risk charts were further recalibrated in 2019 and expanded from 14 WHO regions to 21 global regions, making them more region-specific. For this, data from 85 prospective cohorts with 376,177 individuals from the emerging risk factor collaboration (ERFC) was analyzed and validated and found to be internally and for some countries, externally valid [24,42](Table 3). For internal validation, 19 cohorts that were not included in derivation were used, and the C-statistics found ranged from 0.67 to 0.77 for non-laboratory algorithms, and 0.76 for laboratory algorithms [42]. Risk categories were revised from <10%, 10-<20%, 20-<30%, 30-<40%, and ≥30% in an earlier version to make <5% (green), 5% to <10% (yellow), 10% to <20% (orange), 20% to <30% (red), and ≥30% (dark red) (also, discrete risk range between 1 and 40%) [24].

### ASCVD Risk Score

With changing trends in CVD epidemiology, the American College of Cardiology (ACC) and the American Heart Association (AHA) identified a need to provide an updated and reliable clinical guideline to reduce the risks of atherosclerotic cardiovascular disease (ASCVD) events in different cohorts that can be applied by health care providers. They pooled data from several long-standing population-based cohort studies, such as the Atherosclerosis Risk in Communities (ARIC) study [56], the Cardiovascular Health Study [57], the Coronary Artery Risk Development in Young Adults (CARDIA) study [58], and the Framingham Original and Offspring cohort studies [59,60].

As a result, the pooled cohort equation was derived to provide sex and race-specific estimates of the 10-yr risk for ASCVD for African-American and White men and women 40 to 75 years of age with at least 12 years of follow-up for observation of endpoint occurrence [16]. For this, several novel and classical risk factors were critically evaluated, and their statistical merit, age, total and HDL-cholesterol, systolic BP (irrespective of treatment status), diabetes, and current smoking status were included in the equation. The work group also identified some of the potential risk factors (hs-CRP, CAC, ApoB, albuminuria, GFR, or cardiorespiratory fitness, and ABI); however, due to a lack of adequate evidence, they did not qualify for inclusion statistically. A cut-off of ≥7.5% for 10-year risk estimation was considered to categorize an individual in an elevated (high) risk group. The equation was subjected to internal validation, and the work group also invited additional research to increase its applicability to diversified ethnic groups. In spite of this internal validation exercise and recommendation of current ACC/AHA guidelines for use of pooled cohort scoring for cardiovascular risk assessment, its accuracy for risk prediction is challenged by many ethnic groups, including Americans [61,62].

### QRISK

QRISK is a cardiovascular risk scoring system developed exclusively from the QRESEARCH cohort database (for derivation) and THIN and other large databases (for validation) of patients admitted in different practice centers in the United Kingdom. It was aimed to provide a more accurate risk prediction as compared to the classical Framingham method. The QRISK^27^ was initially developed in 2007 for ethnic groups living in England and Wales and was updated in 2008 and 2017 as the QRISK2 [28] and QRISK3 [29], respectively. Along with classical risk variables from the Framingham equation, the original QRISK model also used additional risk factors for cardiovascular disease, such as deprivation, family history of premature coronary heart disease, body mass index, and the effect of existing antihypertensive treatment. Incorporating population-specific risk factors imparted better discrimination power to QRISK than the Framingham model for the UK population. Immediately after a year in 2008, the original QRISK model was upgraded as QRISK2 to incorporate self-assigned ethnicity and additional risk factors (type 2 diabetes, rheumatoid arthritis, atrial fibrillation, and chronic renal disease (stages 4 and 5) in the risk prediction algorithm. Again, this version was found to have better discrimination statistics than QRISK (with marginal improvement) [28] and the Framingham score [48,63–65]. Importantly, this version noted that ethnicity and deprivation are independent parameters. However, the concern of underestimating CVD risk in patients from deprived areas and overestimating for patients from affluent areas persisted with this algorithm. QRISK3 was published following the NICE on lipid modification and cardiovascular risk assessment that were published in 2014. The guideline highlighted the need to assess and incorporate further potential markers of CVD, in addition to the earlier QRISK2 model, such as HIV/AIDS, stage 3 kidney disease, systemic lupus erythematosus (SLE), severe mental illness, use of atypical antipsychotics or corticosteroids, variation in SBP, and erectile dysfunction, as the exclusion of these conditions may have underestimated the risk in the population requiring preventive intervention. The latest version showed not only higher accuracy and better calibration, but also a large increase in variance, R^2^ (an additional 16% of QRISK2), and the D-statistics (Women and Men, 2.49 and 2.26 in QRISK3, respectively, from 1.80 and 1.62, and 1.78 and 1.60 in QRISK2 and QRISK, respectively) [17,29]. Higher values of these statistics indicate better performances. Despite these positive performance metrics, it is a more resource-demanding algorithm.

### SCORE2/SCORE2-OP

The Systematic Coronary Risk Evaluation (SCORE) project was initiated, and later in 2003, an algorithm was developed, to estimate CVD 10-year risk among Europeans as it differed from that of the FRS [30]. Applying the Weibull proportional hazards model, authors included 205,178 individuals aged 24-68 years from 12 European cohort studies, mainly from general population settings, with good to excellent internal validation (C-statistics, range, 0.74-0.84) [30]. However, the model remained with certain limitations and was also recommended to be revised by the European Society of Cardiology (ESC). The SCORE2 working group initiated to update the model, further, to rectify the age groups of 40-69 (SCORE2) and 70-89 (SCORE2-OP, Older People).

SCORE2 was derived from 677684 individuals (with 30121 CVD events) enrolled from different 45 prospective cohorts in 13 European countries and validated against 1,133,181 individuals from 25 cohorts in 15 countries, for a median duration of 10.7 (5^th^/95^th^ Percentile, 5/18.6) years of follow-up (Table 2) [31]. This algorithm included age, sex, TC, HDL-C, and smoking status and showed good and excellent, internal and external validity, respectively (Tables 1, 4).

**Table 4.**
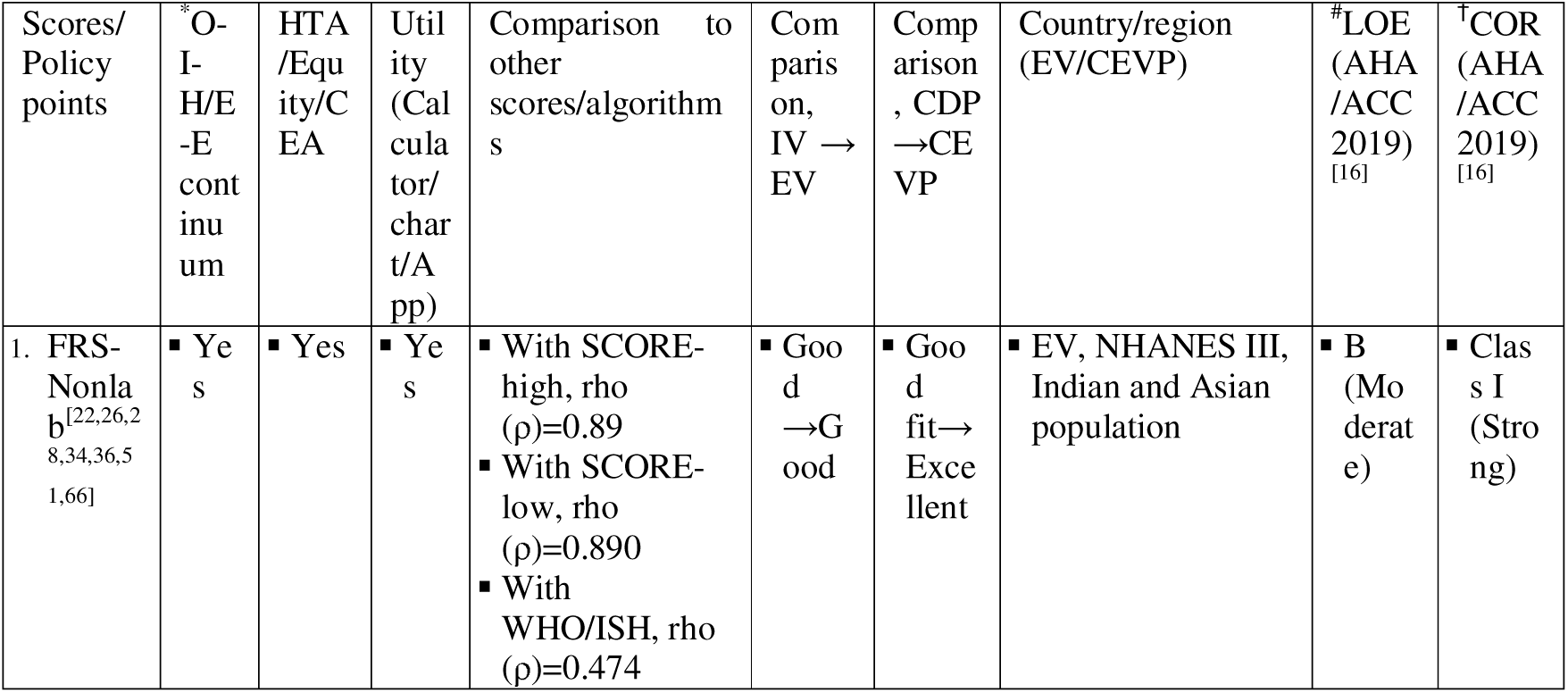
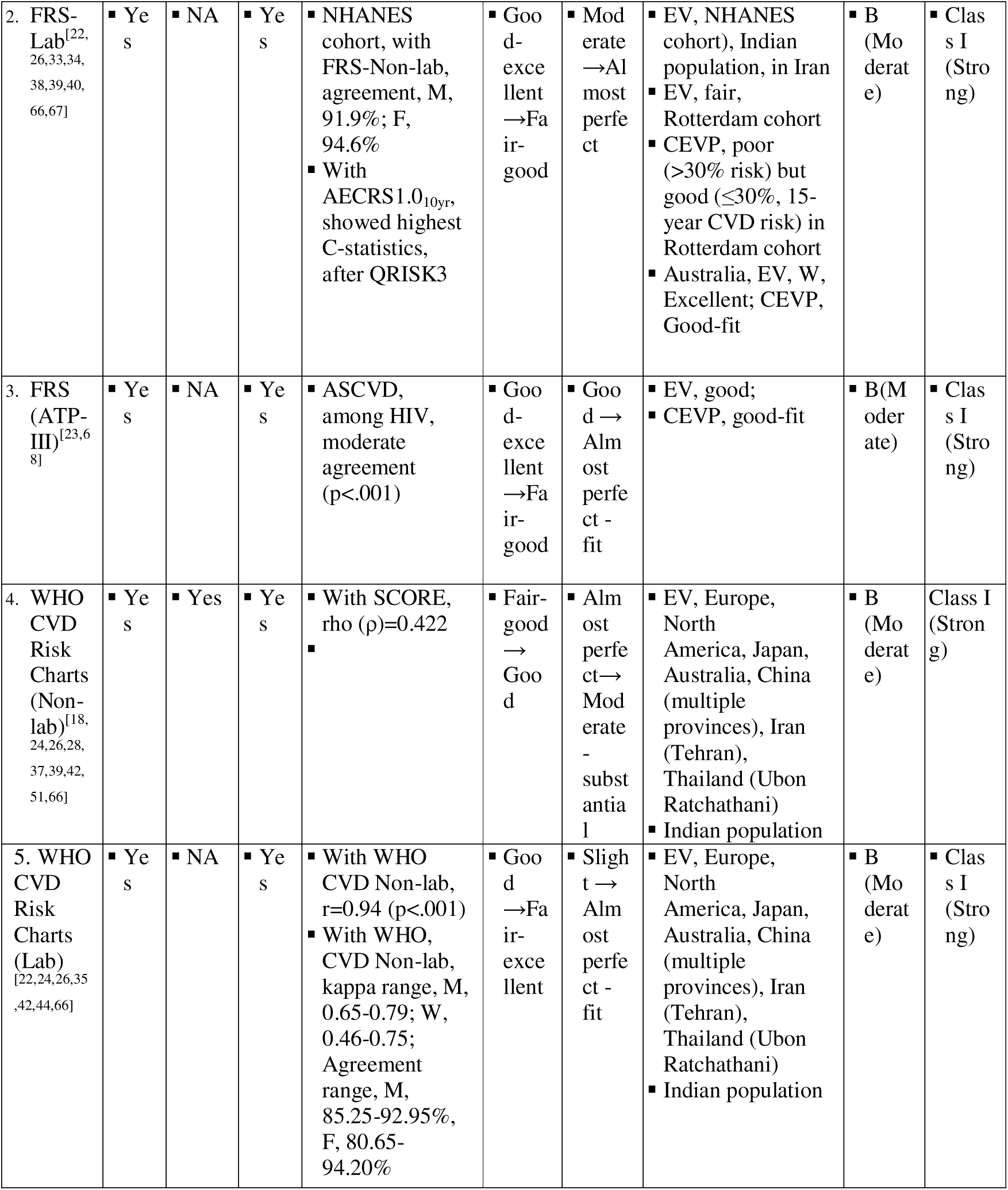
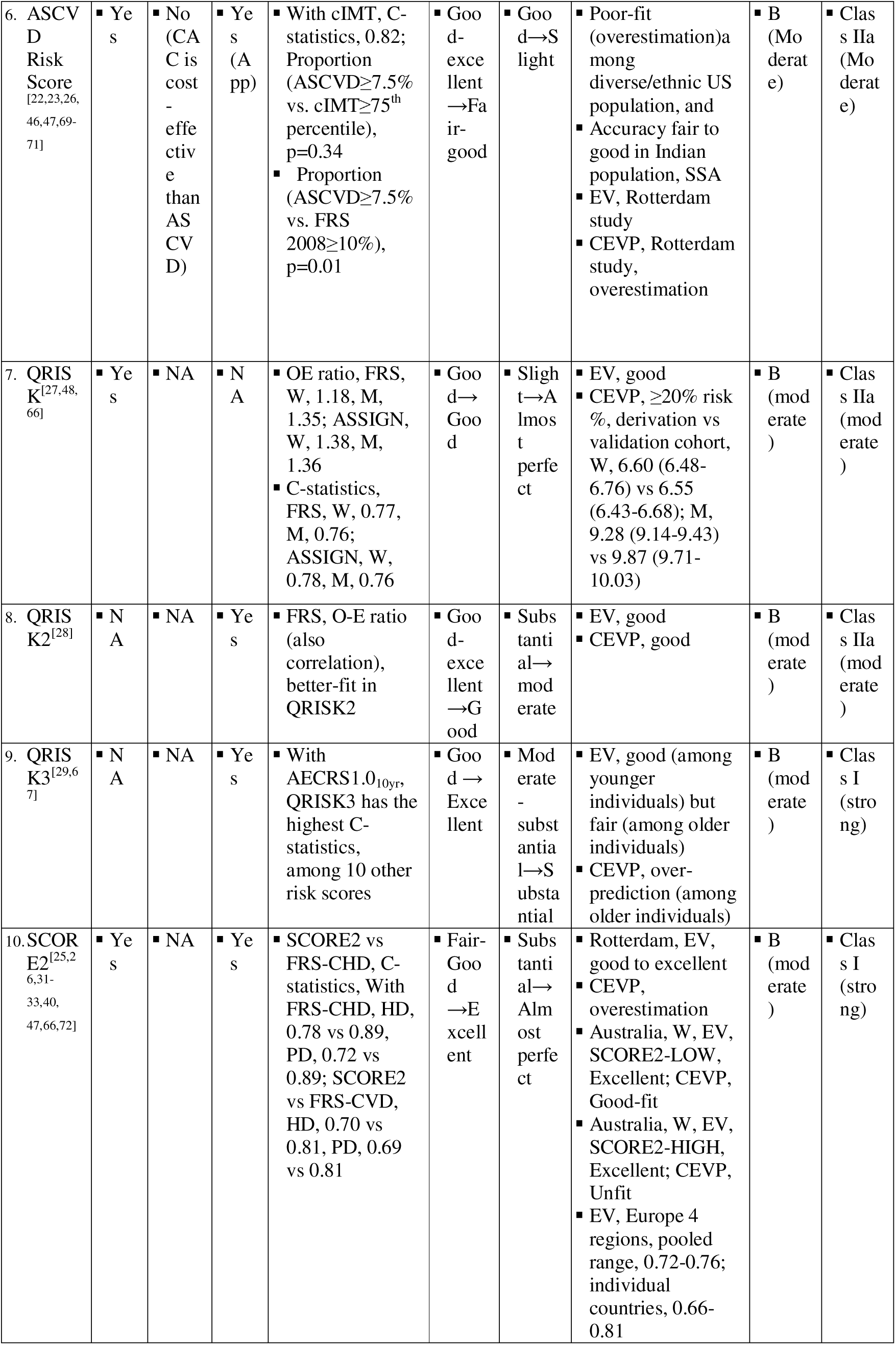
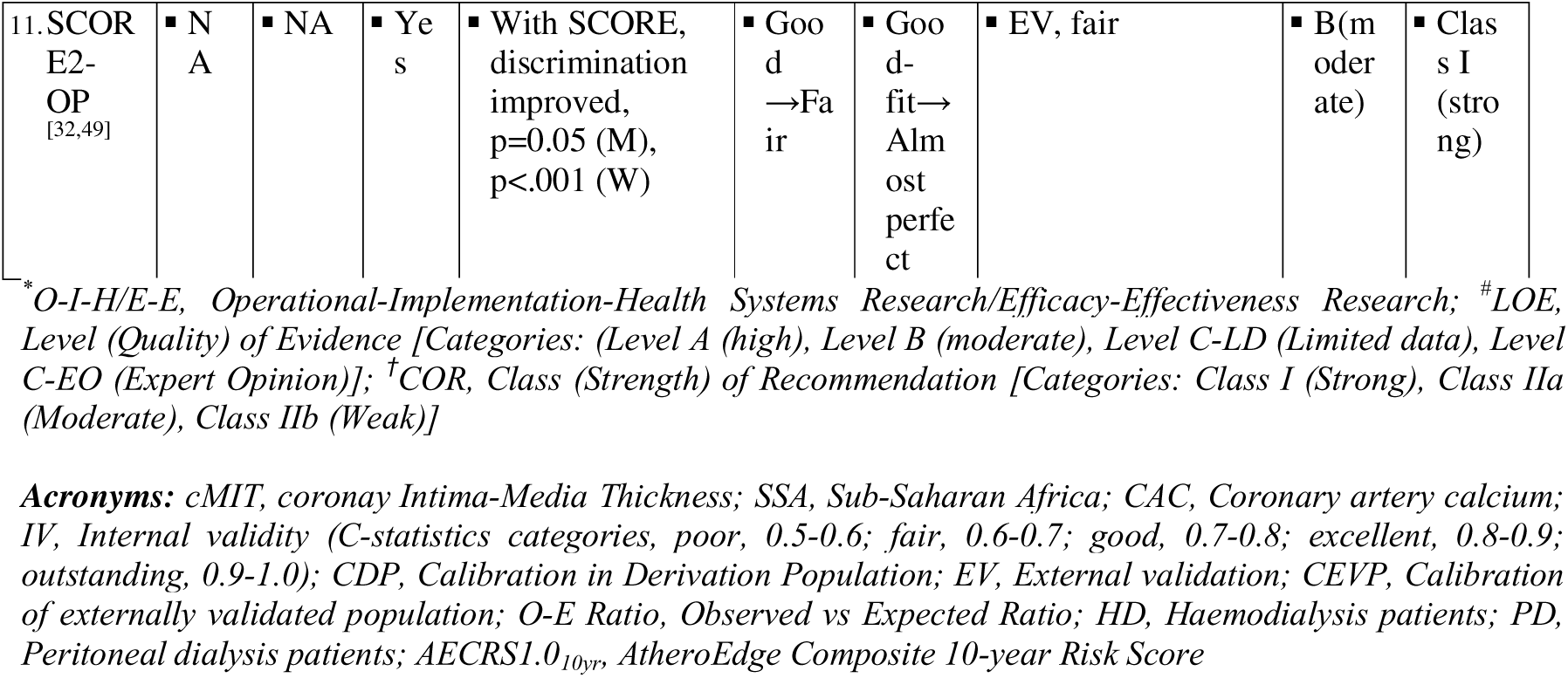
Policy implications of CVD risk scores.

SCORE2-OP was derived from 28,503 participants aged 65 years and older (with 10,089 CVD events) enrolled during 1994-2003 from the Cohort of Norway (CONOR) and externally validated against 338,615 individuals from 6 cohorts (Table 2) for a median duration of 13 (Q1-Q3, 8-15) years of follow up [32]. The algorithm showed good and fair, internal and external validity, respectively, and good fit of predicted vs observed ratio in calibration in the derivation population (Tables 1, 4). A single interactive version to calculate the 10-year CVD risk and the 5- and 10-year CVD risks, for SCORE2 and SCORE2-OP is available as an online calculator, as ‘HeartScore’, and in charts.

## Discussion and prospects

Inclusion of any parameter (especially lab-based) in an algorithm is weighed against the trade-off between disease-mongering and evidence-based health care, including cost [73,74]. Our review revealed that the internal and external validities and the calibrations in the derivation and validation populations not only vary but are also reversed in some cases, both internal and external. Herein, we discuss, first, the gene-environment co-variates (Fig 3.) and, second, the techniques that help predict better. Age, sex, and region are the mostly used covariates, which affected the scores less accurately, so SCORE2 and SCORE2-OP were developed [31,32].

**Fig 3.**
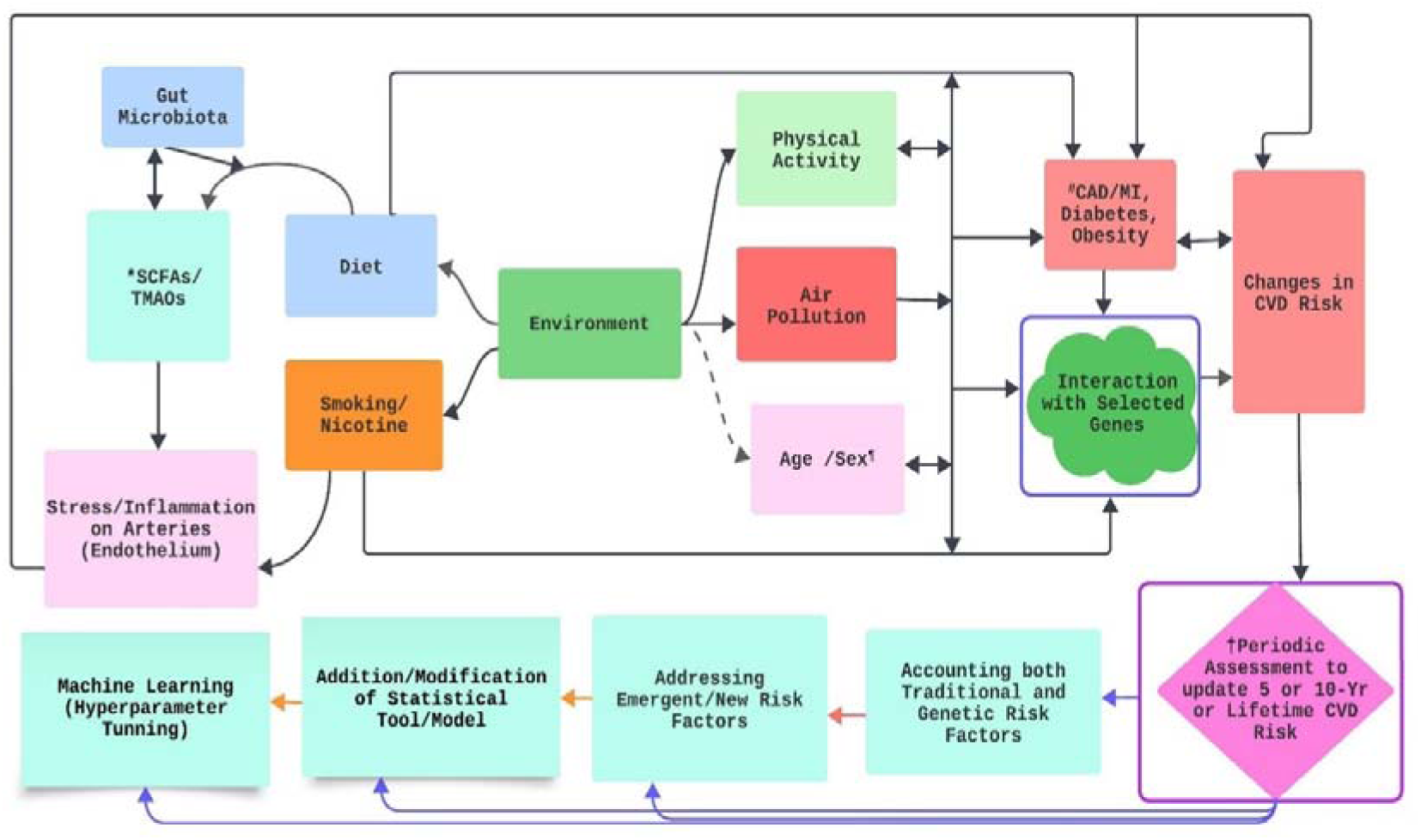
Possible pathways of gene-environment interaction, its impact on CVD Risk, and ways to include in scoring algorithm. The interaction can be manifested, synergized or antagonized. Diet, smoking, physical activities and air pollution have direct interaction with selected genes that are susceptible to CVD risk, as well as through impacting CAD/MI, Diabetes and obesity. Diet, which includes indigestible dietary fiber, which produce SCFAs (having protective effects on CVDs) and red meat and animal proteins, which produce TMAOs (having harmful effects on CVDs), as metabolites, in presence of gut microbiota. Diet may also be enriched in pre-, pro-, and synbiotics, which help further balance the gut microbiota dysbiosis. Smoking and nicotine have also direct effects on endothilium. *SCFAs, Short-chain fatty acids TMAOs, Trimethylamine N-oxides #CAD/MI, Coronary Artery Disease/Myocardial Infarction; ^¶^Environment has indirect effects (dashed line) on age and sex, increasing risk with age in both sexes, and among male until female are menopausal. †Follows updating methods as suggested by Moons et al. (http://dx.doi.org/10.1136/heartjnl-2011-301247), including the given four techniques, solitarily (Indigo lines) or progressive cumulatively (orange lines), in consideration with cost-effectiveness. The periodic time duration depends upon prediction duration, demographic, epidemiological or policy-related factors. One-sided arrow, unidirectional effect: Two-sided arrow, bidirectional effect.

Advanced and modified statistical methods have further improved when recalibrated, both in accuracy and better fit. Sub-distribution hazard ratios (SHRs), which replaced the Weibull model, furthering the F-score along with C-statistics, have shown some improvement [31,44]. The addressal of confounders remained at the center since most of the scores were validated through observational designs, so adjustment models, including the Fine and Gray competing risk adjustment [31]. Moreover, increasing the number of parameters looks cost-effective, but it takes time and increases complexity when applied. A simulation has revealed that a decision tree may be a better option for the k nearest neighbor (kNN) strategy [75].

We propose three ways for furtherance. Firstly, to better capture the variance, rather than limiting with conventional regression models, use of machine learning (ML) with random forest modelling [76], including the Balanced Random Forest algorithm, the Recursive Feature Elimination (RFE) method, and automatic variable selection approach, with inclusion of broader cardiometabolic diseases (CMDs) and other less frequently used diseases such as abdominal aortic aneurism (AAA) [77], are included in the model. Similarly, lasso regression (L1 regularized logistic regression) and the Support Vector Machine (SVM), Regularized Logistic Regression (RLR), and Random Forests (RF) along with a feature selection process carried out with the ML showed the best prediction model in the Xinjiang rural cohort population and the rural Andhra State of India, respectively [44,78]. To add on, a simple statistical calculation, the F-score, can be added to the conventional regression logit for further hierarchical screening [44]. Second, addressing just the “classical risk factors” started in 1948 in Framingham is far from over, and so, now, their “precursors”, which lead to these risk factors, such as carotid plaque composition, which can be assessed through magnetic resonance imaging (carotid MRI), be included in the algorithm [79], which can improve the reclassification of 7.4% and 15.8% with and without cardiovascular events, respectively [80]. In the same vein, coronary artery calcium measurement may help further screen the ‘false negative’ cases in ASCVD [25], whereas pericardial fat volume may be a stronger predictor to substitute the BMI and waist circumference (WC) [81]. On top of this, ‘risk factor to precursor’ shift may help young adults (18-39 age-group), who have not been controlled with ‘risk factor’ approach in the last two decades, as suggested by the National Heart, Lung, and Blood Institute (NHLBI) [82].

Thirdly, rather ‘reinventing the wheel’, we have, to some extent, ‘efficacious’ tools and algorithms such as FRS Non-lab, WHO CVD Risk 2019-Nonlab, which have been found to be cost-effective and more region-specific [42,51] and so, can be re-evaluated using a similar technique for WHO/ISH carried out by Raghu et al in rural Andhra Pradesh of India [44] or external validation, or hyperparameter tuning with sensitive and specific non-invasive risk predictors like waist-hip-ratio (WHR) or WC may still be considered for non-invasive algorithms (Fig 2.), after tested through operational–implementation–health systems and in implementation–effectiveness continuums [82], in many locations and countries that are in need.

While this review provides recent insights of globally used CVD risk algorithms, it is limited by a comprehensive search strategy, some important risk algorithms such as Joint British Society (JBS) Risk Score and its further updated versions, Reynolds Risk Score (RSS), INTERHEART Risk Scores (Fasting Cholesterol and Non-Lab), and others, are beyond the scope of this review. Furthermore, we covered only the findings from selected and recent studies validated and compared with the original ones since it is almost impossible to compile all of them. Also, a clear and scientific alienation between internal and external properties of an algorithm is further demanded, we tried to some extent though.

## Conclusions

For internal and external validation, the corresponding C-statistics for the different risk ratings were 0.65-0.79 and 0.63-0.86, respectively, with given gold standards. However, when consolidated separately for men and women, slightly better ratings were observed for both sexes and only with women, in derivation and validation populations, respectively. In a similar vein, every scoring instrument except ASCVD, QRISK, and QRISK2, which had a moderate strength of recommendation, demonstrated a moderate degree of evidence and was qualified for a strong recommendation (Tables 3, 4).

While the 10-yr CVD risk event prediction accuracy was found to be good to outstanding, there is still room for improvement, with 0.21-0.35 points for derivation and 0.14-0.37 points for validation, for existing gold standards. Risk scores may perform better still if new covariates, mostly “precursors” of risk factors, are included and various statistical techniques, including machine learning, are used (Fig 2.).

## Acronyms and Abbreviations

ASCVD: Atherosclerotic Cardiovascular Disease
CDP: Calibration in Derivation Population
CEVP: Calibration in external validation population
E-E: Efficacy-Effectiveness Research Continuum
EV: External validation
FRS: Framingham Risk Score
IV: Internal validity
O-I-H: Operational-Implementation-Health Systems Research Continuum
QRISK: Cardiovascular Risk Scoring System developed from the QRESEARCH
SCORE: Systematic Coronary Risk Evaluation

## Supporting information

Cover letter

## Data Availability

All data produced in the present work are contained in the manuscript.

## References

1. Benjamin EJ, Blaha MJ, Chiuve SE, et al. Heart disease and stroke statistics—2017 update: A report from the American Heart Association. Circulation. 2017;135(10):e146–603.

2. WHO. Cardiovascular Diseases: Key facts [Internet]. 2021 [cited 2022 Feb 5]. World Health Organization. https://www.who.int/news-room/fact-sheets/CVDS

3. Harikrishnan S, Jeemon P, Mini GK, Thankappan KR, Sylaja P. GBD 2017 Causes of Death Collaborators. Global, regional, and national age-sex-specific mortality for 282 causes of death in 195 countries and territories, 1980-2017: a systematic analysis for the Global Burden of Disease Study 2017. Lancet. 2018 Nov 10;392(10159):1736–1788. 2018;

4. Bennett JE, Kontis V, Mathers CD, et al. NCD Countdown 2030: Pathways to achieving Sustainable Development Goal target 3.4. Lancet. 2020;396(10255):918–34.

5. Bloom DE, Cafiero E, Jané-Llopis E, et al. The global economic burden of noncommunicable diseases. *PGDA Working Papers* 8712. Program on the global demography of aging; 2012.

6. Roth GA, Huffman MD, Moran AE, et al. Global and regional patterns in cardiovascular mortality from 1990 to 2013. Circulation. 2015;132(17):1667–78.

7. Franco M, Cooper RS, Bilal U, Fuster V. Challenges and opportunities for cardiovascular disease prevention. Am J Med. 2011;124(2):95–102.

8. Carnethon MR, Pu J, Howard G, et al. Cardiovascular health in African Americans: a scientific statement from the American Heart Association. Circulation. 2017;136(21):e393–423.

9. Adhikari C, Dhakal R, Adhikari LM, et al. Need for HTA supported risk factor screening for hypertension and diabetes in Nepal: A systematic scoping review. Front Cardiovasc Med. 2022;2090.

10. Chia YC, Gray SYW, Ching SM, Lim HM, Chinna K. Validation of the Framingham general cardiovascular risk score in a multi-ethnic Asian population: a retrospective cohort study. BMJ Open. 2015;5(5):e007324.

11. Selvarajah S, Kaur G, Haniff J, et al. Comparison of the Framingham Risk Score, SCORE and WHO/ISH cardiovascular risk prediction models in an Asian population. Int J Cardiol. 2014;176(1):211–8.

12. Otgontuya D, Oum S, Buckley BS, Bonita R. Assessment of total cardiovascular risk using WHO/ISH risk prediction charts in three low and middle income countries in Asia. BMC Public Health. 2013 Jun 5;13(1):539.

13. Cardiovascular risk prediction tools for populations in Asia. J Epidemiol Community Health. 2007 Feb 1;61(2):115.

14. Findlay SG, Kasliwal RR, Bansal M, Tarique A, Zaman A. A comparison of cardiovascular risk scores in native and migrant South Asian populations. SSM - Popul Health. 2020 Aug 1;11:100594.

15. Hussain SM, Oldenburg B, Wang Y, Zoungas S, Tonkin AM. Assessment of cardiovascular disease risk in South Asian populations. Int J Vasc Med. 2013.

16. Arnett DK, Blumenthal RS, Albert MA, et al. 2019 ACC/AHA Guideline on the Primary Prevention of Cardiovascular Disease: Executive Summary: A Report of the American College of Cardiology/American Heart Association Task Force on Clinical Practice Guidelines. Circulation. 2019 Sep 10;140(11):e563–95.

17. Parsons RE, Liu X, Collister JA, Clifton DA, Cairns BJ, Clifton L. An independent external validation of the QRISK3 cardiovascular risk prediction model applied to UK Biobank participants. medRxiv. 2022 Jan 1;2022.06.30.22277083.

18. Landis JR, Koch GG. The measurement of observer agreement for categorical data. Biometrics. 1977;159–74.

19. Pencina MJ, D’Agostino Sr RB, Demler OV. Novel metrics for evaluating improvement in discrimination: Net reclassification and integrated discrimination improvement for normal variables and nested models. Stat Med. 2012;31(2):101–13.

20. Bitton A, Gaziano T. The Framingham Heart Study’s impact on global risk assessment. Prog Cardiovasc Dis. 2010;53(1):68–78.

21. Mahmood SS, Levy D, Vasan RS, Wang TJ. The Framingham Heart Study and the epidemiology of cardiovascular diseases: A historical perspective. Lancet. 2014;383(9921):999–1008.

22. Garg N, Muduli SK, Kapoor A, et al. Comparison of different cardiovascular risk score calculators for cardiovascular risk prediction and guideline recommended statin uses. Indian Heart J. 2017 Jul 1;69(4):458–63.

23. NHLBI RAWG. Assessing Cardiovascular Risk: Systematic Evidence Review From the Risk Assessment Work Group, 2013 [Internet]. US Department of Health and Human Services, National Institute of Health. www.nhlbi.nih.gov/health-topics/assessing-cardiovascular-risk

24. WHO. Hearts: Technical Package for Cardiovascular Disease Management in Primary Health Care: Risk-based CVD management [Internet]. Geneva: World Health Organization; 2020. https://apps.who.int/iris/handle/10665/333221

25. Lloyd-Jones DM, Braun LT, Ndumele CE, et al. Use of Risk Assessment Tools to Guide Decision-Making in the Primary Prevention of Atherosclerotic Cardiovascular Disease: A Special Report From the American Heart Association and American College of Cardiology. Circulation. 2019 Jun 18;139(25):e1162–77.

26. Durairaj G, Oommen AT, Pillai MG. A cross-sectional validation study comparing the accuracy of different risk scores in assessing the risk of acute coronary syndrome among patients in a tertiary care hospital in Kerala. Indian Heart J. 2020;72(2):113–8.

27. Hippisley-Cox J, Coupland C, Vinogradova Y, Robson J, May M, Brindle P. Derivation and validation of QRISK, a new cardiovascular disease risk score for the United Kingdom: prospective open cohort study. BMJ. 2007;335(7611):136.

28. Hippisley-Cox J, Coupland C, Vinogradova Y, Robson J, Minhas R, Sheikh A, et al. Predicting cardiovascular risk in England and Wales: Prospective derivation and validation of QRISK2. BMJ. 2008;336(7659):1475–82.

29. Hippisley-Cox J, Coupland C, Brindle P. Development and validation of QRISK3 risk prediction algorithms to estimate future risk of cardiovascular disease: prospective cohort study. BMJ. 2017;357.

30. Conroy RM, Pyörälä K, Fitzgerald A el, et al. Estimation of ten-year risk of fatal cardiovascular disease in Europe: the SCORE project. Eur Heart J. 2003;24(11):987–1003.

31. SCORE2 working group and ESC Cardiovascular risk collaboration. SCORE2 risk prediction algorithms: new models to estimate 10-year risk of cardiovascular disease in Europe. Eur Heart J. 2021 Jul 1;42(25):2439–54.

32. SCORE2-OP working group and ESC Cardiovascular risk collaboration. SCORE2-OP risk prediction algorithms: Estimating incident cardiovascular event risk in older persons in four geographical risk regions. Eur Heart J. 2021 Jul 1;42(25):2455–67.

33. Rezaei F, Seif M, Gandomkar A, Fattahi MR, Hasanzadeh J. Agreement between laboratory-based and non-laboratory-based Framingham risk score in Southern Iran. Sci Rep. 2021;11(1):1–8.

34. Pandya A, Weinstein MC, Gaziano TA. A Comparative Assessment of Non-Laboratory-Based Versus Commonly Used Laboratory-Based Cardiovascular Disease Risk Scores in the NHANES III Population. PLOS ONE. 2011 May 31;6(5):e20416.

35. Rezaei F, Seif M, Gandomkar A, et al. Comparison of laboratory-based and non-laboratory-based WHO cardiovascular disease risk charts: a population-based study. J Transl Med. 2022 Mar 16;20(1):133.

36. Goff DC, Lloyd-Jones DM, Bennett G, et al. 2013 ACC/AHA Guideline on the Assessment of Cardiovascular Risk. Circulation. 2014 Jun 24;129(25_suppl_2):S49–73.

37. Schnabel RB, Sullivan LM, Levy D, et al. Development of a risk score for atrial fibrillation (Framingham Heart Study): A community-based cohort study. Lancet. 2009;373(9665):739–45.

38. D’Agostino RB, Pencina MJ, Massaro JM, Coady S. Cardiovascular Disease Risk Assessment: Insights from Framingham. Global Heart. 2013 Mar 1;8(1):11–23.

39. van Kempen BJ, Ferket BS, Kavousi M, et al. Performance of Framingham cardiovascular disease (CVD) predictions in the Rotterdam Study taking into account competing risks and disentangling CVD into coronary heart disease (CHD) and stroke. Int J Cardiol. 2014;171(3):413–8.

40. Goh LGH, Welborn TA, Dhaliwal SS. Independent external validation of cardiovascular disease mortality in women utilising Framingham and SCORE risk models: a mortality follow-up study. BMC Women’s Health. 2014;14:1–11.

41. Cooney MT, Dudina AL, Graham IM. Value and limitations of existing scores for the assessment of cardiovascular risk: a review for clinicians. J Am Coll Cardiol. 2009;54(14):1209–27.

42. Kaptoge S, Pennells L, De Bacquer D, et al. WHO cardiovascular disease risk charts: Revised models to estimate risk in 21 global regions. Lancet Glob Health. 2019;7(10):e1332–45.

43. WHO. Prevention of cardiovascular disease: Guidelines for assessment and management of total cardiovascular risk. Prev Cardiovasc Dis Guidel Assess Manag Cardiovasc Risk [Internet]. 2007. https://apps.who.int/iris/handle/10665/43685

44. Raghu A, Praveen D, Peiris D, Tarassenko L, Clifford G. Implications of Cardiovascular Disease Risk Assessment Using the WHO/ISH Risk Prediction Charts in Rural India. PLOS ONE. 2015 Aug 19;10(8):e0133618.

45. Chia YC, Lim HM, Ching SM. Validation of the pooled cohort risk score in an Asian population–a retrospective cohort study. BMC Cardiovasc Disord. 2014;14:1–7.

46. Rana JS, Tabada GH, Solomon MD, et al. Accuracy of the atherosclerotic cardiovascular risk equation in a large contemporary, multi-ethnic population. J Am Coll Cardiol. 2016;67(18):2118–30.

47. Kavousi M, Leening MJG, Nanchen D, et al. Comparison of Application of the ACC/AHA Guidelines, Adult Treatment Panel III Guidelines, and European Society of Cardiology Guidelines for Cardiovascular Disease Prevention in a European Cohort. JAMA. 2014 Apr 9;311(14):1416–23.

48. Hippisley-Cox J, Coupland C, Vinogradova Y, Robson J, Brindle P. Performance of the QRISK cardiovascular risk prediction algorithm in an independent UK sample of patients from general practice: a validation study. Heart. 2008;94(1):34–9.

49. Cooney MT, Selmer R, Lindman A, et al. Cardiovascular risk estimation in older persons: SCORE OP. Eur J Prev Cardiol. 2016;23(10):1093–103.

50. Verweij L, Peters RJG, Scholte op Reimer WJM, et al. Validation of the Systematic COronary Risk Evaluation - Older Persons (SCORE-OP) in the EPIC-Norfolk prospective population study. Int J Cardiol. 2019 Oct 15;293:226–30.

51. Pandya A, Weinstein MC, Salomon JA, Cutler D, Gaziano TA. Who Needs Laboratories and Who Needs Statins? Comparative and Cost-Effectiveness Analyses of Non–Laboratory-Based, Laboratory-Based, and Staged Primary Cardiovascular Disease Screening Guidelines. Circ Cardiovasc Qual Outcomes. 2014;7(1):25–32.

52. Wilson PWF, D’Agostino RB, Levy D, Belanger AM, Silbershatz H, Kannel WB. Prediction of coronary heart disease using risk factor categories. Circulation. 1998;97(18):1837–47.

53. D’Agostino RB, Grundy S, Sullivan LM, Wilson P. Validation of the Framingham coronary heart disease prediction scores: Results of a multiple ethnic groups investigation. JAMA. 2001;286(2):180–7.

54. D’Agostino Sr RB, Vasan RS, Pencina MJ, et al. General cardiovascular risk profile for use in primary care: the Framingham Heart Study. Circulation. 2008;117(6):743–53.

55. Abidov A, Chehab O. Cardiovascular risk assessment models: Have we found the perfect solution yet? J Nucl Cardiol. 2020;27.

56. ARIC Investigators. The atherosclerosis risk in communit (ARIC) study: Design and objectives. Am J Epidemiol. 1989;129(4):687–702.

57. Fried LP, Borhani NO, Enright P, et al. The cardiovascular health study: Design and rationale. Ann Epidemiol. 1991;1(3):263–76.

58. Friedman GD, Cutter GR, Donahue RP, et al. CARDIA: study design, recruitment, and some characteristics of the examined subjects. J Clin Epidemiol. 1988;41(11):1105–16.

59. Dawber TR, Kannel WB, Lyell LP. An approach to longitudinal studies in a community: The Framingham Study. Ann N Y Acad Sci. 1963 May 22;107:539–56.

60. Kannel WB, Feinleib M, McNamara PM, Garrison RJ, Castelli WP. An investigation of coronary heart disease in families. The Framingham offspring study. Am J Epidemiol. 1979 Sep;110(3):281–90.

61. Ridker PM, Cook NR. Statins: New American guidelines for prevention of cardiovascular disease. Lancet. 2013;382(9907):1762–5.

62. Amin NP, Martin SS, Blaha MJ, Nasir K, Blumenthal RS, Michos ED. Headed in the right direction but at risk for miscalculation: A critical appraisal of the 2013 ACC/AHA risk assessment guidelines. J Am Coll Cardiol. 2014;63(25 Part A):2789–94.

63. Collins GS, Altman DG. An independent external validation and evaluation of QRISK cardiovascular risk prediction: A prospective open cohort study. BMJ. 2009;339.

64. Collins GS, Altman DG. An independent and external validation of QRISK2 cardiovascular disease risk score: A prospective open cohort study. BMJ. 2010;340.

65. Collins GS, Altman DG. Predicting the 10-year risk of cardiovascular disease in the United Kingdom: Independent and external validation of an updated version of QRISK2. BMJ. 2012;344.

66. Muthee TB, Kimathi D, Richards GC, et al. Factors influencing the implementation of cardiovascular risk scoring in primary care: A mixed-method systematic review. Implement Sci. 2020 Jul 20;15(1):57.

67. Khanna NN, Jamthikar AD, Gupta D, et al. Performance evaluation of 10-year ultrasound image-based stroke/cardiovascular (CV) risk calculator by comparing against ten conventional CV risk calculators: A diabetic study. Comput Biol Med. 2019 Feb 1;105:125–43.

68. Elsamadisi P, Cha A, Kim E, Latif S. Statin use with the ATP III guidelines compared to the 2013 ACC/AHA guidelines in HIV primary care patients. J Pharm Pract. 2017 Feb 1;30(1):64–9.

69. Muhlestein JB., Knowlton KU, Le VT, et al. Coronary artery calcium versus pooled cohort equations score for primary prevention guidance. JACC Cardiovasc Imaging. 2022 May 1;15(5):843–55.

70. Gluckman TJ, Kovacs RJ, Stone NJ, Damalas D, Mullen JB, Oetgen WJ. The ASCVD risk estimator app. J Am Coll Cardiol. 2016 Jan 26;67(3):350–2.

71. Mosepele M, Hemphill LC, Palai T, et al. Cardiovascular disease risk prediction by the American College of Cardiology (ACC)/American Heart Association (AHA) Atherosclerotic Cardiovascular Disease (ASCVD) risk score among HIV-infected patients in sub-Saharan Africa. PLOS ONE. 2017 Feb 24;12(2):e0172897.

72. Gabriella AA, Stanescu C, Burcea AF, et al. MO760: Cardiovascular score risk calculators in dialysis patients: Can we use them? Nephrol Dial Transplant. 2022 May 1;37(Supplement 3).

73. Bonner C, McKinn S, McCaffery K, et al. 23 Heart age: intuitive communication tool or gateway to overdiagnosis? A comparison of 3 countries. BMJ Evid-Based Med. 2019 Dec 1;24(Suppl 2):A18.

74. Reynolds T. Targeting treatment using calculated cardiovascular disease risk: Effective? A shot in the dark? Or is it time to change the paradigm? Int J Clin Pract. 2009;63(12):1785–91.

75. Bhowmick P, Gajjar S, Chaudhary S. Hyperparameter tuning and comparison of k nearest neighbour and decision tree algorithms for cardiovascular disease prediction. Int J Swarm Intell. 2021;6(2):118–29.

76. Yang L, Wu H, Jin X, et al. Study of cardiovascular disease prediction model based on random forest in eastern China. Sci Rep. 2020 Mar 23;10(1):5245.

77. Rezaee M, Putrenko I, Takeh A, Ganna A, Ingelsson E. Development and validation of risk prediction models for multiple cardiovascular diseases and Type 2 diabetes. PLOS ONE. 2020 Jul 29;15(7):e0235758.

78. Qian X, Li Y, Zhang X, et al. A cardiovascular disease prediction model based on routine physical examination indicators using machine learning methods: A cohort study. Front Cardiovasc Med. 2022;9.

79. Saba PS, Parodi G, Ganau A. From Risk Factors to Clinical Disease. J Am Coll Cardiol. 2021 Mar 23;77(11):1436–8.

80. Zavodni AE, Wasserman BA, McClelland RL, et al. Carotid artery plaque morphology and composition in relation to incident cardiovascular events: The multi-ethnic study of atherosclerosis (MESA). Radiology. 2014;271(2):381–9.

81. Miao C, Chen S, Ding J, et al. The association of pericardial fat with coronary artery plaque index at MR imaging: The multi-ethnic study of atherosclerosis (MESA). Radiology. 2011;261(1):109–15.

82. Gooding HC, Gidding SS, Moran AE, et al. Challenges and opportunities for the prevention and treatment of cardiovascular disease among young adults: Report from a national heart, lung, and blood institute working group. J Am Heart Assoc. 2020;9(19):e016115.

