## Supplemental Table 1 for "Appraising Cardiovascular 10-yr Risk Prediction Scores: A Rapid Systematic Review"

SM1.Tab1. Artificial Intelligence guided search for five prioritized CVD 10-year risk scores

| SN | Three AI Inputs, Outputs and Synthesis of Five CVD Risk Scores in Order | | | | | Final Selection |
|---|---|---|---|---|---|---|
| | *Input | ChatGPT | Perplexity | Gemini | Synthesis | |
| 1 | Five **most valid** CVD 10-year risk scores or algorithms | FRS, ASCVD, QRISK3, SCORE2, RSS | FRS, SCORE, ASCVD, WHO, QRISK3 | PCE, ASCVD, ASCVD for Women, FRS, RRS | FRS, ASCVD, QRISK, SCORE, RRS | FRS, ASCVD, SCORE, QRISK, #WHO 2019 |
| 2 | Five **globally optimally used** CVD 10-year risk scores or algorithms | FRS, ASCVD, QRISK3, SCORE2, RSS | FRS, SCORE, ASCVD, QRISK, WHO | FRS, PCE, RSS, ASCVD, ASCVD for Women | FRS, ASCVD, QRISK, SCORE, RRS | |
| 3 | Five CVD 10-year risk scores or algorithms with **high utilities for resource-constraint settings** | WHO, INTERHEART, FRS-Simplified Version, SCORE-Cholesterol-Free Version, ASCVD-Modified for Low-Resource Setting | WHO/ISH, WHO-Non-Lab, SCORE, FRS, Globorisk | FRS, RRS, PCE, Simplified PCE, Clinical Risk Stratification Tools | FRS, WHO, SCORE, ASCVD/PCE | |

**Predicate in bold used to emphasize the search in AI; #Consensused by two review authors (CA and KS) to select WHO instead of RSS to represent high utility in resource-constraint settings; ChatGPT, Chat Generative Pre-trained Transformer; CVD, Cardiovascular diseases; QRISK, QRESEARCH Cohort Database; WHO 2019, World Health Organization CVD Risk 2019; INTERHEART, INTERHEART Modifiable Risk Score; RRS, Reynolds Risk Score; SCORE, Systematic Coronary Risk Evaluation; ASCVD/PCE, Atherosclerotic Cardiovascular Disease (ASCVD) Risk Estimator/Pooled Cohort Equation; ISH, International Society of Hypertension*
