## Supplementary material for "Appraising Cardiovascular 10-yr Risk Prediction Scores: A Rapid Systematic Review": Cover letter

Date: 8^th^ Feb, 2025

To

The medXiv Team

Re: **Submission of a manuscript**

Dear Team,

Greetings from Pokhara University!!

We would like to submit a review manuscript entitled “Appraising Cardiovascular 10-yr Risk Prediction Scores: A Rapid Systematic Review” for your kind consideration. This review aims to summarize methodological, statistical, and implementational aspects of globally used cardiovascular risk prediction scores, and also discuss how they can be used by different countries and states, mainly those who have not validated yet, for ad hoc, and for, future strategies to develop. To the best of our knowledge no updated review or consensus is available regarding this. Moreover, among the Asians, being house of phenotypically and genotypically diversified population along with numerous LMICs need contextual scoring systems for cardiovascular diseases. In addition, the review is being authored by researchers from India and Nepal, conditioned and expert (also working with authors from other Asian countries) and hence provide a good perspective for Asians and similar LMICs. Some authors are also part of HTAIn (Health Technology Assessment India) regional resource center and hence have provided critical aspect of cost in the review which might be useful for LMICs (figure 1). So, the review may be helpful for program designers, implementors, and policy makers. We registered a protocol in the Open Science Framework (<https://osf.io/72v48>) before carrying out the review.

Our systematic but restricted search confining five risk scores (and their derivations), and synthesis, revealed many of the globally used risk scores are not validated, mainly in LMICs, nor there are consistencies between and among the external accuracies with that of internal. Moreover, search also emphasizes the imperativeness of either calibration of existing risk scores with the primary data or develop and validate new ones. Herein, we propose and discuss how these available risk algorithms can be utilized ad hoc and for long-term policies.

All of the authors meet the ICMJE criteria for authorship and we declare that we are keeping global and/or local guidelines of research and publication ethics including authorship.

Since this study involves a review of previously published literature and does not involve direct interaction with human subjects or the collection of new data, ethical approval is not required.

Kindly let me know if any further information is required.

With warm regards,

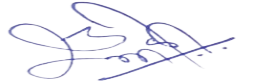

Chiranjivi Adhikari, MPH, MED, PhD Candidate

Indian Institute of Public Health Gandhinagar (IIPHG), IN/Pokhara University, NP

Cell/Whatsapp: +977-9819046979 (NP)

Corresponding Author
